# Pupil size-resting state network coupling in major depressive disorder: a longitudinal iTBS study

**DOI:** 10.64898/2026.10.27.26366619

**Authors:** Monica Di Giuliano, Andy Schumann, Martin Walter, Carsten Schmidt-Samoa, Roberto Goya-Maldonado

## Abstract

Altered central-peripheral autonomic nervous system communication plays a key role in major depressive disorder (MDD). In this context, pupillary dynamics may be considered a non-invasive peripheral proxy of treatment-related changes during intermittent theta-burst stimulation (iTBS) in MDD. However, the relationship between spontaneous pupillary fluctuations and the resting-state functional networks (RSNs) involved in MDD is still underexplored. Here, we investigated longitudinal changes in pupil–RSN coupling in patients with MDD undergoing an iTBS intervention. Resting-state functional magnetic resonance imaging with concurrent pupillometry was acquired at four study visits. Group independent component analysis was used to estimate tonic pupil coupling with predefined RSNs of interest, along with a linear mixed-effects model to assess longitudinal changes in pupil size–RSN coupling and, exploratively, the effect of potential clinical and behavioral moderators. Among the examined RSNs, the left executive control network (lECN), including the left dorsolateral prefrontal cortex as the cortical target of stimulation, exhibited a sustained reduction in positive pupil size coupling relative to baseline, irrespective of treatment order (active - sham). To determine whether this longitudinal change was accompanied by a more general change in pupil diameter, we additionally examined pupil size independently of RSNs. Pupil diameter decreased significantly from baseline, with greater decreases being positively associated with greater reductions in anxiety scores. Notably, reduction in anxiety symptoms were also positively correlated with reduction in depressive symptoms. Together, these findings suggest that longitudinal reductions in pupil-IECN coupling occur alongside reductions in pupil diameter that are associated with reductions in anxiety symptoms.

## 1 INTRODUCTION

Major depressive disorder (MDD) is a common psychiatric disorder, characterized by depressed mood, loss of interest or pleasure, and other symptoms that impair daily functioning, including feelings of guilt or worthlessness, disturbed sleep or appetite, fatigue and difficulties with concentration and memory (Valenza, 2023). Diagnostic criteria define depression by the presence of a specified number of depressive symptoms and functional impairment contributing to the assessment of clinical severity and illness burden (American Psychiatric Association, 2013; Zimmerman et al., 2018). MDD is also considered a major contributor to global disability, with substantial prevalence worldwide (Friedrich, 2017; World Health Organisation, 2026). Pharmacological approaches including antidepressants as well as psychotherapeutical interventions, such as cognitive behavioral therapy, are considered first-line treatment options for MDD. Nonetheless, substantial inter-individual variability in treatment response remains a major challenge, possibly reflecting the clinical and biological heterogeneity of the disorder (Neuteboom et al., 2023; Rush et al., 2006). Accordingly, MDD is characterized by a considerable variation in symptom profiles, severity, onset, and clinical course (Rush et al., 2006; Valenza, 2023).

Non-invasive brain stimulation such as transcranial magnetic stimulation (TMS) have increasingly emerged as potential treatment approaches of MDD (Blumberger et al., 2018; Chung et al., 2015; Citrenbaum et al., 2023; Kishi et al., 2024). TMS involves the application of magnetic field changes through the intact scalp, inducing electric fields in underlying brain tissue and thereby modulating cortical activity (Chung et al., 2015; Kobayashi & Pascual-Leone, 2003). When delivered repetitively, it is referred to as repetitive TMS (rTMS), a protocol that can induce lasting changes in cortical excitability and network function (rTMS; Allen et al., 2007; Chung et al., 2015; Kobayashi & Pascual-Leone, 2003; Maeda et al., 2000). Among the rTMS protocols, theta-burst stimulation (TBS) consists of repeated bursts of the pulses at 50Hz, with bursts repeated at 5 Hz, corresponding to the theta range. Accordingly, intermittent TBS (iTBS), in which these bursts are delivered intermittently, has emerged as an efficient rTMS protocol for the treatment of MDD (Chung et al., 2015; Kishi et al., 2024). Specifically, iTBS has the main advantage of shorter treatment sessions with comparable antidepressant effects (Di Lazzaro et al., 2008; Kishi et al., 2024; Neuteboom et al., 2023; Rounis & Huang, 2020). In MDD, a common target of stimulation is the left dorsolateral prefrontal cortex (dlPFC). In this regard, the dlPFC - a region involved in cognitive control processes and a key hub of the executive control network (ECN) - has been shown to be as effective as some first-line antidepressant treatments (Kishi et al., 2024; Neuteboom et al., 2023; Schiena et al., 2021; Tura et al., 2024). Together with the dlPFC, different studies have outlined the potential adaptive effects of iTBS on different functional and structural brain signatures that characterise the psychopathological profile of MDD, which could be assessed using functional magnetic resonance imaging (fMRI) methods (Schiena et al., 2021).

In this context, pupillary dynamics have emerged as a promising, non-invasive physiological measure that may provide complementary information on arousal and autonomic related processes along rTMS interventions (Citrenbaum et al., 2023; M. Leuchter et al., 2025; M. K. Leuchter et al., 2026). Specifically, pupillary responses reflect processes ranging from sensory regulation and alerting to orienting, arousal, and higher-order cognitive control (Strauch et al., 2022). These responses involve distributed neural systems, including the locus coeruleus-noradrenergic system (LC-NE) and superior colliculus (SC), which in turn interact with sensory and higher-order executive brain regions to shape pupillary dynamics across resting-state and task-related conditions (Aston-Jones & Cohen, 2005; Strauch et al., 2022; C.-A. Wang & Munoz, 2015). Notably, pupillary responses have been associated with fMRI blood oxygenation level-dependent (BOLD) signals in the LC-NE and other cortical and subcortical regions during resting state conditions (DiNuzzo et al., 2019; Schneider et al., 2016). Accordingly, spontaneous low-frequency fluctuations in pupil diameter have been linked to intrinsic brain activity, suggesting that tonic pupillary dynamics may provide a peripheral correlate of ongoing arousal-related and neuromodulatory states both in healthy and in clinical populations (Chang et al., 2013; DiNuzzo et al., 2019; Liu et al., 2026, 2026; Niehaus et al., 2001; Phylactou et al., 2026; Schneider et al., 2016; Weijs et al., 2025; Yellin et al., 2015). For example, in healthy populations, spontaneous pupil fluctuations have been linked to distinct neural signatures across different resting-state functional networks (RSNs). Among these RSNs, the ECN regions, including the dlPFC, are considered crucial giving their role in exerting top-down control over pupillary responses together with the LC-NE and SC systems (Citrenbaum et al., 2023; DiNuzzo et al., 2019; M. K. Leuchter et al., 2026; Strauch et al., 2022). Along with the ECN, pupil linked dynamics have also been associated with fluctuations in multiple other RSNs, including default mode (DMN), salience (SN), and attention networks (DiNuzzo et al., 2019; Ishida et al., 2020; Prompiengchai & Dunlop, 2025; Schiena et al., 2021; Schneider et al., 2016; Yellin et al., 2015). These large-scale networks are also involved in regulating various cognitive, emotional and arousal-related processes that are relevant to disorder psychopathology (Schiena et al., 2021). Crucially, these processes could, in turn, be investigated through tonic pupillary response dynamics (FERENCOVA et al., 2021; Schiena et al., 2021). For instance, tonic pupil size has been found to be associated with individual differences in various cognitive and affective processes related to depression, such as anxiety. Notably, anxiety is considered an affective state that can influence the recruitment of cognitive control resources, thereby affecting the activation of neural correlates underlying these mechanisms, such as the dlPFC, in MDD (FERENCOVA et al., 2021; Fietz et al., 2024; Garner, 2026; Jones et al., 2015; Kahneman & Beatty, 1966; Siegle, Steinhauer, Carter, et al., 2003; Siegle et al., 2004; van der Wel & van Steenbergen, 2018). Thus, overall, pupillary responses may inform of a complementary peripheral measure for investigating processes relevant to the clinical heterogeneity of MDD, considering the symptom levels (Alvares et al., 2016; Aminihajibashi et al., 2019; Citrenbaum et al., 2023; Hu et al., 2016; Hughes & Stoney, 2000; Jones et al., 2015; Phylactou et al., 2026; Schiena et al., 2021; Schumann et al., 2017; Siegle et al., 2004).

It is worth noticing that, in the context of depression, relatively few studies have investigated pupillary responses in relation to rTMS treatment: accordingly, the available evidence suggests that treatment-related changes occurred in terms of an overall increase in adaptive parasympathetic activity, as indexed by changes in pupil diameter and related constriction metrics, over the treatment period (Citrenbaum et al., 2023; M. K. Leuchter et al., 2026). However, to the best of our knowledge, studies investigating pupillary measures in MDD and in the context of TMS have primarily focused on other dimensions of pupillary responses sensitive to autonomic dysfunctions. This includes the pupillary light reflex, which measures reactivity to a brief light stimulus, and measures of pupil constriction and reactivity that may provide information on autonomic function in MDD (Citrenbaum et al., 2023; M. Leuchter et al., 2025; M. K. Leuchter et al., 2026). Nevertheless, the longitudinal relationship between spontaneous low-frequency tonic pupil fluctuations and resting-state functional network activity has not been directly investigated in MDD during an rTMS intervention: this might represent an important gap, as pupil–RSN coupling may provide complementary information on the relationship between peripheral arousal-related dynamics and intrinsic large-scale brain network organisation during treatment (Liu et al., 2026; Schneider et al., 2016; Yellin et al., 2015). To this end, the current study aimed to investigate whether tonic low-frequency fluctuations in pupil diameter are associated with longitudinal changes in coupling with major RSNs during iTBS treatment in MDD. We employed a longitudinal, counterbalanced iTBS design over a 6-week treatment period, combining repeated resting-state fMRI with concurrent pupillometry. We further explored whether treatment order and clinical factors, including depressive and anxiety symptoms, were associated with longitudinal changes in pupil dynamics and pupil–RSN coupling (Aminihajibashi et al., 2019; Jones et al., 2015; Phylactou et al., 2026; Schiena et al., 2021; Siegle et al., 2004).

At the end, by integrating longitudinal pupillometry with long-scale resting state networks, this study ultimately sought to characterise changes in brain–body coupling during iTBS treatment by leveraging tonic low-frequency pupil–RSN dynamics as potential proxy of treatment-related changes in MDD.

## 2 MATERIALS AND METHOD

### 2.1 Participants

The study included 88 individuals aged 18 to 64 years, diagnosed by trained psychiatrists who confirmed the eligibility using the Structured Clinical Interview for DSM-5 Disorders – Clinical Version (SCID-5-CV) and the MADRS score (Hengartner et al., 2020; Montgomery & Åsberg, 1979). Severity of depression was assessed by trained clinical interviewers weekly: notably, responder status to treatment was defined as ≥50% reduction in MADRS total score considering all visits. Intellectual functioning was estimated with the Mehrfach-Wortschatz-Test B (MWTB; Neumann & Wolfram, 1978). To further characterise the clinical profile of the sample, total scores on the Beck Anxiety Inventory (BAI) were examined (Beck et al., 1988; Geissner & Huetteroth, 2018). Exclusion criteria encompassed a history of neurological or other significant psychiatric disorders (except anxiety), contraindications for rTMS or fMRI (e.g., epilepsy, neurological diseases, pregnancy, metallic implants), and changes in antidepressant medication or dosage within the 2 weeks preceding enrollment. Participants receiving psychotropic medications were required to maintain stable medication regimens for a minimum of two weeks prior to and throughout the duration of the study, with adherence monitored through serum level assessments. MRI, blood and urine laboratory assessments were used to exclude organic causes of depressive symptomatology. Out of 88 patients, six subjects were excluded because they met criteria for bipolar disorder rather than MDD.

The study protocol complied with the Declaration of Helsinki and was approved by the Ethics Committee of the University Medical Center Göttingen (UMG). Prior to enrollment, all participants provided verbal and written informed consent after the study protocol was fully explained. For further details on the study design and protocol, see Wilkening et al., 2022.

### 2.2 Study design

The study protocol used a randomized, sham-controlled, quadruple-blinded (participants, care provider, clinical interviewers, and principal investigator), counterbalanced crossover design over six weeks. Each participant received 20 sessions of iTBS per week: one week of active stimulation and one week of sham stimulation. The order of stimulation was randomly assigned (active-sham, or sham-active). Randomization was based on pre-coded sequences generated using a true random function in MATLAB and were managed by the principal investigator, who had no direct contact with the study participants.

This study formed part of a larger project that examines the effects of stimulation site localization within the left dlPFC (Wilkening, Jungeblut, Adamovic, Belov, Dechent, Eicke, Hansen, Kozyrev, Marten, et al., 2025). Thus, two stimulation site approaches were applied: one using the F3 position from the international 10-20 electroencephalography system, and the other using an individualized target based on each participant’s resting-state functional magnetic resonance imaging (rs-fMRI) data. Specifically, the individualized approach selected a position to stimulate the left dlPFC at which the participant exhibited the strongest anticorrelation with the DMN (Singh et al., 2019, 2020; Wilkening, Jungeblut, Adamovic, Belov, Dechent, Eicke, Hansen, Kozyrev, Muschke, et al., 2025). Given that previous analyses found no significant differences in clinical effectiveness between the two approaches, data from both approaches were analyzed together (e.g., Wilkening et al., 2022, 2025). Four MRI scans were conducted at specific time points: before (V1) and after (V2) the first week of stimulation, and before (V3) and after (V4) the second week of stimulation.

The current study design is summarized in **Figure 1**.

**Figure 1.**
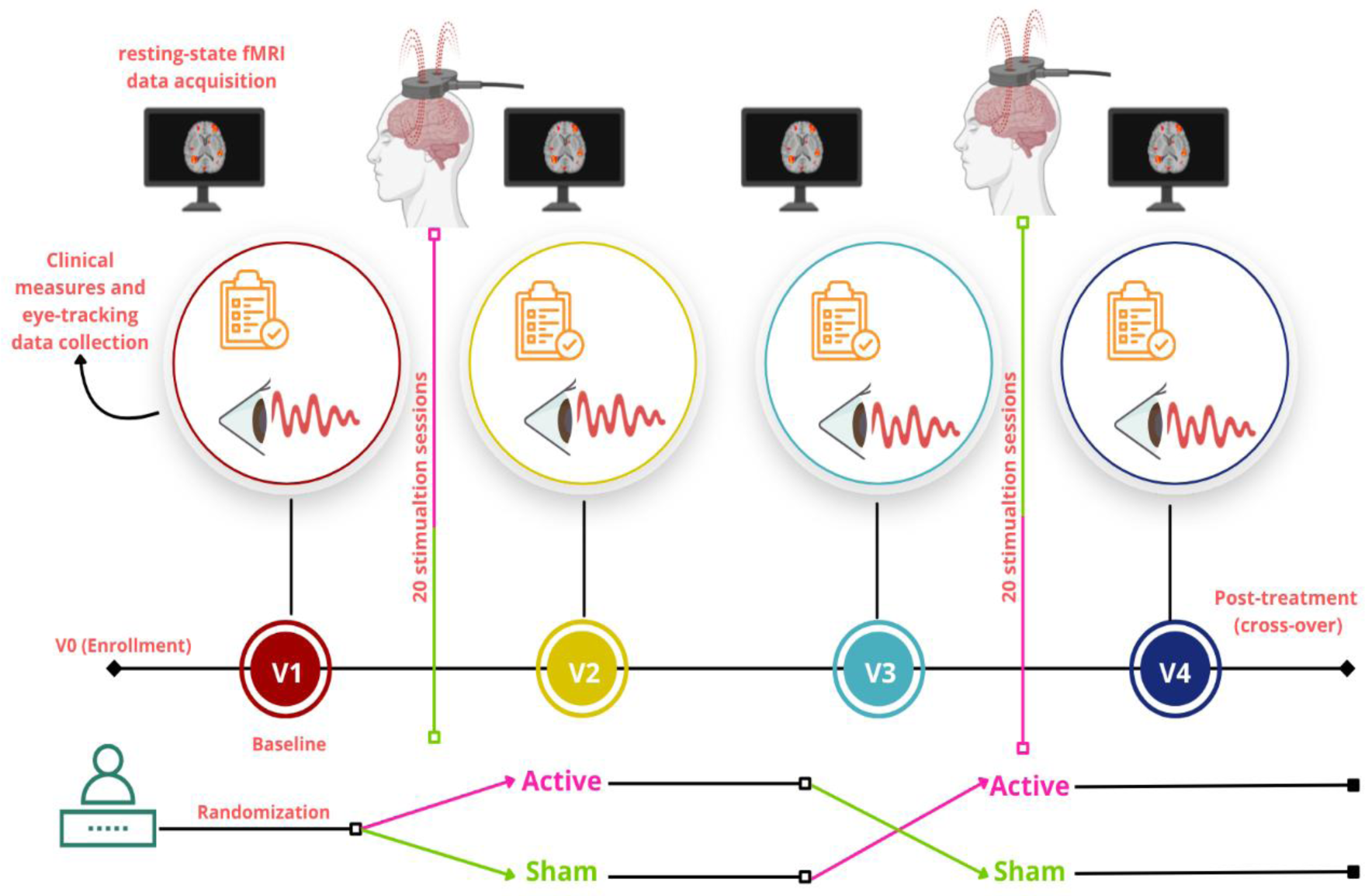
Study design. Overview of the intermittent theta-burst stimulation (iTBS) study design. A schematic representation of the study design, including randomization between active-sham or sham-active iTBS conditions, the four visits (V1 to V4) at which clinical measures including Montgomery-Åsberg Depression Rating Scale (MADRS) and Beck Anxiety Inventory (BAI), resting-state functional MRI as well as concurrent pupillometry data were acquired for subsequent analyses. This figure was created using BioRender (BioRender.com).

### 2.2 iTBS protocol

The MagVenture X100 system with Mag-option and a figure-of-eight MCF-B65 A/P cooled coil was used for stimulation by trained and experienced personnel. Each daily visit included four sessions of approximately 10 min each, followed by at least 20 min pauses, based on the inter-session time described in a previous aftereffect study (Tse et al., 2018). The iTBS protocol consisted of bursts at 5 Hz, each comprising 3 pulses delivered at 50 Hz, resulting in a volley with 2 s on and 8 s off. One session consisted of 60 volleys, resulting in a total of 1800 pulses per session, 7200 per day, and 36000 pulses in total. The resting motor threshold (RMT), defined as the minimum stimulation intensity required to elicit a motor evoked potential (MEP), was reevaluated daily via electromyography (EMG) in the right first dorsal interosseous muscle as the lowest intensity eliciting a motor response of 50 µV in at least 5 of 10 attempts. Treatment stimulation was performed at 110% of the RMT (Goldsworthy et al., 2016). In the sham condition, the same figure-eight-shaped MCF-B65 A/P coil was blindly flipped 180 degrees according to pre-coded sequences. Additionally, transcutaneous electrical nerve stimulation (tENS) electrodes mimicked the scalp sensation simultaneously with the sound of magnetic pulses, regardless of the stimulation condition. The principal investigator predetermined the codes for active and sham protocols to ensure quadruple-blinding. The success of blinding was assessed using a visual analogue scale (VAS), which assessed pain and participants’ expected therapeutic outcomes on a scale of 0 to 10. More detailed descriptions are provided in Wilkening and colleagues (Wilkening et al., 2022).

### 2.3 MRI data acquisition

MRI data were collected on a 3T whole-body system equipped with a 64-element head/neck matrix coil (MAGNETOM Prisma fit, Siemens Healthcare, Erlangen, Germany). During scanning, participants were instructed to keep their heads still and relax without falling asleep or engaging in deliberate, systematic thinking, in a dimly lit room. T2*-weighted images were acquired using a multiband (MB)-accelerated echo-planar imaging (EPI) sequence with a repetition time (TR) of 1.5 s, echo time (TE) of 30 ms, flip angle (FA) of 77°, and a MB acceleration factor of 3. The acquisition used a 104 × 104 matrix and a field of view (FOV) of 210 × 210 mm², resulting in an in-plane resolution of approximately 2 × 2 mm², and 69 transverse slices with a slice thickness of 2 mm and slice spacings of 2.2 mm. Phase encoding was performed in the posterior-to-anterior (PA) direction. The pixel bandwidth was 1550 Hz, and the effective echo spacing was 0.365 ms. Each resting-state fMRI run comprised 400 whole-brain volumes, corresponding to an acquisition duration of approximately 10 min. In addition to functional data, we also acquired high-resolution anatomical T1-weighted volume scans using a magnetization-prepared rapid gradient-echo (MPRAGE) sequence. The MPRAGE parameters were: TR = 2.25 s, TE = 3.3 ms, inversion time (TI) = 900 ms, FA = 9°, matrix size = 256 x 256, voxel size = 1 x 1 x 1 mm, and comprised 176 sagittal slices. Two participants were excluded from further analyses because of incomplete MRI data.

### 2.4 Pupillometry data acquisition and analysis

Eye-tracking of the right eye was performed using a monocular Arrington Research ViewPoint eye-tracking system (ViewPoint PC-60; Arrington Research, Scottsdale, AZ, USA). The MR-compatible 60-Hz MRC camera (model 12M; MRC Systems, Heidelberg, Germany) and infrared light source were mounted on top of the MR head coil, behind a semi-transparent mirror through which participants viewed a screen positioned at the end of the scanner bore. During the resting-state scan, participants fixated on a white cross presented at the center of a dark screen using a Projector ProPIXX system (VPixx Technologies, Saint-Bruno, QC, Canada; 1920 × 1080 pixels; display size: 43 × 24 cm; viewing distance: 89 cm). Horizontal and vertical gaze coordinates and pupil measurements were acquired by the ViewPoint system and synchronized with the experimental presentation and scanner acquisition via Presentation software (Neurobehavioral Systems, Berkeley, CA, USA). Pupil measures were recorded in arbitrary units (AU), as provided by the eye-tracking system, at a nominal sampling rate of 60 Hz. At this stage, a total number of twelve subjects were excluded from further preprocessing because of missing pupil data.

Data were preprocessed following standard procedures used in previous pupillometry studies (Fink et al., 2023; Kret & Sjak-Shie, 2019; Mathôt & Vilotijević, 2023; Schneider et al., 2016; Steinhauer et al., 2022; Yellin et al., 2015). First, blink-related samples were identified using complementary run-specific criteria. Specifically, in addition to a conventional threshold on pupil values falling below the run-specific median minus 2.5 standard deviation, a lower 0.1th-percentile threshold was applied to the pupil distribution to identify the lower boundary (“cliff”) of valid tracking data, thereby reducing the influence of run-to-run differences in blink frequency. Additionally, rapid changes in pupil metrics were identified using a robust velocity criterion based on the median and median absolute deviation (MAD) of the absolute frame-to-frame pupil velocity, normalized by the sampling interval (Kret & Sjak-Shie, 2019; Mathôt & Vilotijević, 2023). This latter criterion was used to ultimately identify abrupt changes in pupil signal that may reflect tracking artifacts associated with rapid eyelid closure or reopening rather than physiological pupil dynamics (Kret & Sjak-Shie, 2019). Velocity outliers were used to expand the initial blink mask only when they coincided with samples already identified by the initial blink criteria, rather than serving as an independent criterion for blink detection. Thus, a blink mask was created by the conjunction of both standard deviation and cliff criteria, expanded when a velocity outlier coincided with this mask and finally padded by 150 ms before and after the occurrence of blink-related events (Kret & Sjak-Shie, 2019; Mathôt & Vilotijević, 2023; Steinhauer et al., 2022). Horizontal and vertical gaze coordinates were screened independently for extreme values the specified number of MADs from their respective medians. Samples identified by either the blink or gaze-artifact mask were removed from the pupil signal. The remaining pupil-related signal was then smoothed using a sample moving-median filter (Kret & Sjak-Shie, 2019; Mathôt & Vilotijević, 2023). Additionally, to reduce pupil-foreshortening error (PFE) associated with gaze position, pupil measurements were corrected using a multiple linear regression model fitted separately for each recording run, with standardized horizontal and vertical gaze coordinates as predictors and an intercept. The estimated contributions of both gaze coordinates were subtracted from the measured pupil, while retaining the intercept (Mathôt & Vilotijević, 2023).

After the first step of cleaning the data for potential blink-related artifacts and gaze-related issues, missing pupil samples were linearly interpolated for gaps of up to 0.5 s, whereas longer gaps were retained as missing for subsequent quality-control assessment (Fink et al., 2023). Specifically, runs were excluded from further analysis if, after interpolation, they contained a maximum remaining gap >8 s, >30% missing data, >25 remaining gaps longer than 1 s, or a cumulative duration of gaps longer than 1 s exceeding 45 s. These exclusion criteria were defined based on both the characteristics of the present recordings, including the eye-tracker sampling rate (60 Hz) and the fMRI repetition time (TR = 1.5 s), and consideration of established approaches for identifying and excluding recordings with excessive missing or artifact-contaminated pupil data (Fink et al., 2023; Steinhauer et al., 2022). In addition to these recording-level exclusion criteria, the longitudinal design of the study was also considered in the subsequent fMRI analyses by including only participants with reliable pupil metrics at both baseline (V1) and the treatment endpoint (V4) in the final analysis sample. Following cleaning and application of the quality control assessment criteria, a total number of 23 participants were excluded from further pre-processing steps as not meeting the inclusion criteria for reliable pupil metrics.

For retained runs, the pupil signal was band-pass filtered between 0.01 and 0.1 Hz using a second-order Butterworth filter with zero-phase filtering. This frequency range was selected based on previous pupillometry and resting-state fMRI studies investigating the spectral characteristics of pupil and BOLD fluctuations and their cross-spectral relationship, while attenuating very slow signal drifts and higher-frequency fluctuations (e.g., Schneider et al., 2016; Yellin et al., 2015). The filtered signal was subsequently downsampled to match the fMRI repetition time (TR = 1.5 s) and normalized. A fixed 1-s forward temporal shift was then applied to the normalized pupil time course by interpolating the signal at time points 1 s later than each fMRI acquisition time. This shift was intended to account for the delayed pupillary response relative to changes in luminance and attentional or arousal-related processes, consistent with previous pupillometry studies (Schneider et al., 2016; Wierda et al., 2012; Yellin et al., 2015). The resulting temporally shifted, downsampled pupil-size signal was used as the main regressor of interest in the first-level general linear model (GLM). The regressor was convolved with the canonical hemodynamic response function (HRF), and the corresponding temporal and dispersion derivatives were included to allow for variability in the latency and temporal dispersion of the BOLD response, respectively (Schneider et al., 2016). For each fMRI volume, the proportion of pupil samples affected by residual blink- or artifact-related gaps was calculated and entered into the first-level GLM as a nuisance parametric regressor. This was intended to account for variance in the BOLD signal associated with temporal variability in pupil-data availability and signal quality, rather than to serve as a pure blink regressor. Thus, the pupil-size effect was estimated conditional on the variance accounted for by this nuisance regressor to reduce confounding of the pupil-size effect by variability in eye-tracking data quality, while acknowledging that blink-related variance may remain in periods where pupil data were successfully interpolated (Schneider et al., 2016). A better description of the GLM model is provided in the Method section 2.6.

Finally, the blink- and artifact-cleaned, interpolated pupil data from the retained visits of the final sample were additionally used to further characterize changes in tonic pupil size across the course of treatment, independently of their association with resting-state brain fluctuations. To this end, we calculated the mean pupil diameter for each visit and tested longitudinal changes using a linear mixed-effects model (Section 2.8).

Pupillometry preprocessing steps can be schematically seen in **Figure 2**.

**Figure 2.**
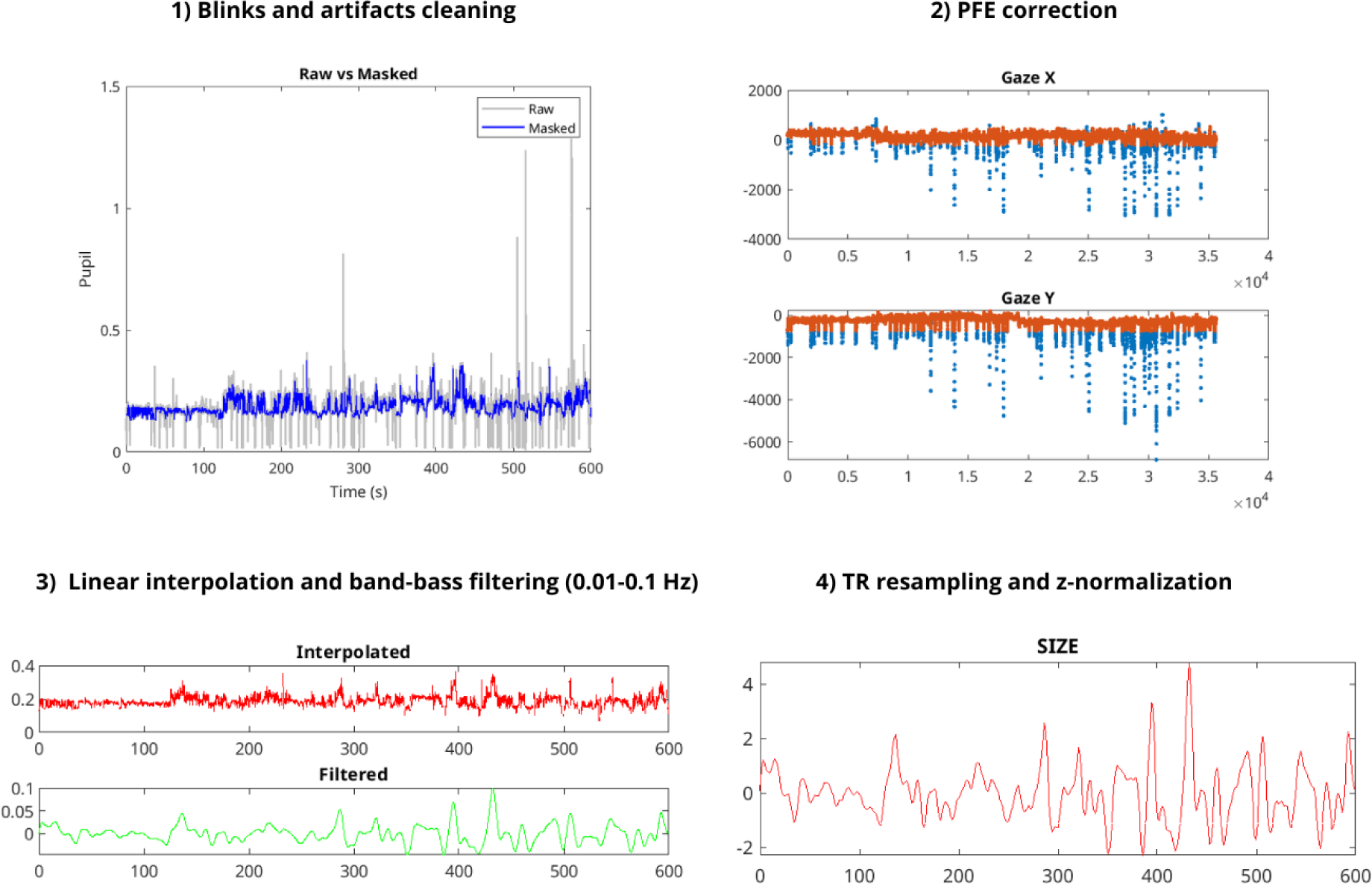
Overview of the main pupillometry preprocessing steps. An example of the pupil time course recorded from a single participant during resting-state fMRI. The preprocessing pipeline comprised: (1) generation of a combined artifact mask based on predefined criteria for blinks and other signal artifacts; (2) correction for pupil-foreshortening error (PFE) to reduce the potential influence of gaze position on the measured pupil signal, in conjunction with the removal of blink-related artifacts; (3) linear interpolation of short data gaps followed by band-pass filtering (0.01–0.1 Hz) to isolate spontaneous low-frequency pupil oscillations during rest; and (4) resampling of the cleaned pupil signal to the fMRI repetition time (TR), followed by z-score normalization, to obtain the tonic pupil-size regressor used in subsequent analyses. Pupil measurements are expressed in arbitrary units (AU) throughout the preprocessing steps; the final pupil-size regressor is expressed as a z-scored measure.

### 2.5 rsfMRI preprocessing

Data analysis was performed using SPM12 (https://www.fil.ion.ucl.ac.uk/spm). The preprocessing pipeline included slice-timing correction, realignment, gradient echo field map unwarping, coregistration to the anatomical scan, segmentation, and spatial normalization to the Montreal Neurological Institute (MNI152) template.

Nuisance signals from white matter (WM) and cerebrospinal fluid (CSF) were extracted using subject-specific tissue probability maps generated during SPM’s unified segmentation–normalization procedure: WM probability maps were thresholded at 99% probability and binarized. To further reduce the contamination by adjacent gray matter, the resulting WM masks were subjected to two iterations of binary morphological erosion and subsequently restricted to the functional brain mask (Muschelli et al., 2014). CSF probability maps were thresholded at 70% probability and binarized, then constrained to the ventricular regions defined by the ALVIN mask and subsequently restricted to the functional brain mask (Kempton et al., 2011; Muschelli, 2020; Muschelli et al., 2014). An anatomical component-based noise correction (aCompCor) was performed on the voxelwise fMRI time series extracted from the resulting WM and CSF masks. This procedure involved a linear detrending of the time series and mean-centering prior to principal component analysis (PCA; (Behzadi et al., 2007). PCA was performed using singular value decomposition, and the first five principal components from each tissue compartment were retained as nuisance regressors. Head-motion parameters were modeled using a 24-parameter Friston model comprising the six rigid-body motion parameters, their temporal derivatives, and the squared terms of both the original parameters and their derivatives. Framewise displacement (FD) was calculated from the six rigid-body motion parameters using a 50-mm head-radius approximation (Power et al., 2012). Global DVARS and gray-matter-restricted DVARS - calculated within a gray-matter mask with a probability threshold of 40% - were also computed to quantify changes in the fMRI signal (Muschelli et al., 2014). For quality-control purposes, volumes were flagged as outliers when FD exceeded 0.5 mm or when gray-matter DVARS exceeded 1.5 standard deviations above its run-specific mean (Muschelli et al., 2014; Power et al., 2012a). Visits with >30% of volumes exceeding the FD threshold were excluded from further fMRI analyses. This additional FD-based exclusion criterion was intended to identify visits with excessive frame-to-frame displacement, which may introduce transient motion-related signal changes not adequately addressed by continuous regression of motion parameters (Power et al., 2012). On this basis, a total number of 4 participants were excluded from further analyses because they exceeded the FD-based exclusion threshold. Finally, a voxelwise nuisance regression was performed separately for each visit. The regression model included the 24 motion parameters, the five WM aCompCor components, and the five CSF aCompCor components (Behzadi et al., 2007; Muschelli et al., 2014; Van Schuerbeek et al., 2022). Following regression, the original temporal mean of each voxel was restored to the residual time series before reconstruction of the cleaned 4D fMRI dataset. The resulting functional images in MNI space were then smoothed using a Gaussian kernel with 6-mm full width at half maximum (FWHM) to improve the signal-to-noise ratio.

A first-level GLM was then specified with a high-pass filter cut-off of 250 s, an AR(1) model of serial correlations, and the canonical hemodynamic response function including temporal and dispersion derivatives. Pupil-related regressors were entered as parametric modulators of the volume events (later removed from the model), with the proportion of missing pupil data entered first as a nuisance parametric modulator and pupil size entered second as the regressor of interest. Accordingly, a parametric modulation approach adapted to resting state data was used to orthogonalize the pupil-size regressor with respect to the missing-data regressor. This meant that the estimate of the pupil-size regressor could reflect the component of pupil-size variation that was independent of missing-data-related variance (Schneider et al., 2016). For each retained subject-specific visit, the FD-based volume-specific nuisance regressors were also included into the first-level GLM together with the pupil-related regressors. The pupil-size regressor estimated parametric modulation was then extracted from each subject- and visit-specific first-level SPM design matrix and used for the temporal regression analysis of the spatial group Independent Component Analysis (ICA), to estimate the association between pupil-related fluctuations and the time course of each RSNs (ICA) component (Section 2.7 below).

### 2.6 Spatial ICA analysis

Spatial ICA was performed on the entire data set using the ‘‘Group ICA for fMRI Toolbox’’ (GIFT; http://mialab.mrn.org/software/gift), implemented in MATLAB. Given the unbalanced nature of the final sample and the subsequent longitudinal second-level analyses of pupil-related fluctuations, each subject-specific visit (V1, V2, V3, and V4) was treated as an independent dataset for the group spatial ICA. The ICA was estimated jointly across all subject-visit datasets using the default GIFT algorithms, allowing the identification of common spatially independent components across visits while retaining subject- and visit-specific component estimates for subsequent analyses. The Minimum Description Length (MDL) criterion was used for model-order selection to reduce potential selection bias and estimate the dimensionality of the signal subspace in fMRI data (V. D. Calhoun & De Lacy, 2017). Specifically, MDL selects a model that balances data approximation with model complexity (V. D. Calhoun & De Lacy, 2017). The number of independent components (ICs) was set to a standard number of 20. Twenty to 50 components have been commonly used in fMRI studies: higher numbers of components generally yield more spatially focal components, whereas lower numbers tend to capture broader networks (V. D. Calhoun & De Lacy, 2017; Di Giuliano et al., 2025). To address the randomness of the initialization inherent in ICA decomposition, we used ICASSO in conjunction with the Infomax algorithm. ICASSO was used to quantitatively assess the stability of the ICA estimates across repeated turns with different random initializations (Himberg et al., 2004). Infomax, in turn, is an ICA algorithm that estimates independent components by maximizing mutual information (V. D. Calhoun & De Lacy, 2017; Himberg et al., 2004; Li et al., 2007). Data were reduced through two principal component analysis (PCA) stages (first subject-specific PCA = 25; second group-based PCA = 20) and concatenated at each stage for further reduction. The concatenated data were then used for group ICA estimation and back-reconstruction of component time courses and spatial maps for each participant (group-ICA/GICA back reconstruction step). The resulting single-subject time course amplitudes were then calibrated (scaled) using the raw data in order to reflect the percentage of fMRI signal change for comparison across participants. The individual back-reconstructed components were then used to compute mean and standard deviation spatial maps and time courses, as well as t-statistic/z-shifted spatial maps (V. D. Calhoun et al., 2001; V. D. Calhoun & De Lacy, 2017).

We used the spatial and temporal regression procedures implemented in GIFT to identify the RSNs of interest across visits and estimate their temporal associations with pupil size for subsequent second-level analyses. Specifically, for spatial identification, the 20 independent components were labelled using a canonical resting-state network template (NeuroMark 2.2 template) provided by GIFT. Given the aims and design of the study, we focused on the left- and right-lateralised executive control networks (ECNs), which are implicated in executive control, playing a role in modulating pupillary dynamics, and encompass stimulation target, as well as six other RSN networks of interest: the visual network, DMN (dorsal and ventral), the SN, and the dorsal and ventral attention networks (DAN+VAN). Following visual inspection to exclude noise-related components, we retained components based on their relevance to spontaneous pupil-related fluctuations and their coupling with BOLD fluctuations, particularly in the context of depression (Jensen et al., 2024; Prompiengchai & Dunlop, 2025; Razi et al., 2017; Schiena et al., 2021; Schneider et al., 2016; Strauch et al., 2022; S. Wang et al., 2026; Yellin et al., 2015). Spatial z-scored maps of the selected seven networks - identified as common RSNs across all visits - are presented in the Supplementary Materials **(Fig. S1)** in sagittal, coronal, and axial views.

Finally, the temporal regression algorithm implemented in the GIFT toolbox was adapted to quantify the association between pupil-related fluctuations and the time courses of the selected ICs. For each subject and visit, the first-level SPM design matrix was used to identify the parametric modulation corresponding to pupil size (i.e., the canonical hemodynamic response function-convolved pupil size regressor). Specifically, this subject- and visit-specific regressor was extracted directly from the estimated first-level design matrix and used as the primary regressor of interest in the temporal regression analysis, while the remaining regressors from the corresponding first-level design matrix were retained as nuisance/control regressors. For each IC, its GIFT-derived time course was entered as the dependent variable in a multiple linear regression model, with the pupil-size regressor as the predictor of interest and the remaining first-level regressors as covariates. This approach was used to estimate the pupil-specific regression coefficient (β), representing the direction and magnitude of the association between pupil size and IC activity after accounting for the other regressors, for each IC and subject-visit dataset. The resulting subject- and visit-specific β estimates were subsequently used as the dependent variable in the longitudinal linear mixed-effects analysis (LMEs; section 2.8).

### 2.7 Statistical analyses

Longitudinal changes in pupil-derived physiological measures (mean pupil diameter) and pupil size–ICs beta estimates were assessed using a LMEs framework, with a participant-specific random intercept to account for repeated observations. The model was estimated using restricted maximum likelihood (REML). Age (mean-centered) and sex were included as covariates in each LME model.

For the primary longitudinal analyses, treatment visits (V1, V2, V3, and V4) were modeled as a categorical within-subject factor, with intervention sequence (active-sham vs. sham-active iTBS) included as a between-subject factor. The longitudinal design was interpreted according to treatment sequence: for participants starting with either the active or sham condition, V1 represented baseline (pre-treatment), V2 the acute post-intervention assessment, V3 the latest intervention assessment before crossover, and V4 the post-crossover assessment. The primary analysis tested the longitudinal effects of iTBS on pupil size coupling with each of the selected RSNs (ICs) by testing the main effect of Visit, while accounting for intervention order as fixed effect. P-values were corrected for multiple comparisons using the false discovery rate (FDR). Separate exploratory linear mixed-effects models (LMEs) were conducted for the primary pupil size–RSN beta estimates and mean pupil diameter to investigate (i) the Visit × intervention-sequence interaction (active–sham vs. sham–active iTBS), (ii) differences according to MADRS-defined response status, coded categorically as responders and non-responders (see section 2.1), and (iii) associations with anxiety symptoms, assessed using total BAI scores. Each model additionally included the corresponding lower-order main effects of intervention sequence and the relevant clinical measures, including MADRS and BAI.

Additionally, we performed exploratory analyses restricted to the baseline (V1) and end-of-treatment (V4) time points to assess whether individual changes in MADRS total scores and BAI total scores were associated with changes in mean pupil diameter or changes in the primary pupil size-IC beta estimates. First, we performed paired-samples t-tests to assess within-subject changes in physiological and clinical data between the time-matched baseline (V1) and post-treatment (V4) visits, with corresponding Cohen’s (d_z_) effect sizes calculated. Then, to examine the associations between changes in the primary pupil–IC beta findings as well as mean pupil diameter measures with changes in BAI/MADRS total scores, change scores (Δ = V4 − V1) were calculated using a Pearson correlation analysis.

Finally, for findings considered for interpretation in the main Results, sensitivity analyses were performed to assess the influence of individual participants or observations. Depending on the analysis structure, these included leave-one-subject-out analyses for participant-level associations and leave-one-observation-out analyses for longitudinal mixed-effects models, together with inspection of standardized residuals, consistent with established approaches to influence assessment in longitudinal mixed-effects models (Zhang & Hedeker, 2023).

## 3 RESULTS

### 3.1 Sample characteristics

A final sample of 41 participants with unipolar MDD (mean age = 32.5 ± 12.1) was analyzed based on the quality control and inclusion criteria established to ensure reliable pupil measures at both V1 and V4 (Section 2.5) as well as MRI data (Section 2.4). Among these 41 participants, 27 had reliable physiological and MRI data available for analysis at V2, 29 at V3, and 21 had complete data across all four visits. The mean age at the primary diagnosis of MDD was 19.5 (± 8.9). Additionally, 3 of the 41 participants were left-handed.

The sample was characterized by a premorbid verbal intelligence estimate (IQ) of 29.9 ± 2.9, as measured using the MWBT. Regarding the clinical characteristics of the sample (Table 1), depressive symptoms decreased significantly from V1 (24.07 ± 8.2) to V4 (18.60 ± 8.8), as measured using the MADRS (p < 0.001; Cohen’s d_z_ = −0.68). Furthermore, 28 of the 41 participants were classified as non-responders (section 2.1). Anxiety symptoms, as assessed by total BAI scores, also decrease significantly from V1 (18.29 ± 10.3) to V4 (9.53 ± 7.1), with p < 0.001 (Cohen’s d_z_ = −1.11). The individual trajectories for MADRS and BAI total scores are presented in **Fig. 3A and 3B**. We also found a significant positive correlation between changes in BAI and MADRS total scores, indicating that greater reductions in anxiety symptoms were associated with greater reductions in depressive symptoms from baseline to post-treatment (r = 0.46, p = 0.002; see **Fig.4**). The association between changes in BAI and MADRS scores remained robust when potential influential observations were considered, as no participant showed an extreme change score and leave-one-participant-out analyses yielded significant correlation (r = 0.40 - 0.52, all p ≤ 0.01). Finally, the physiological data that mean pupil diameter significantly decreased from V1 (0.18 AU ± 0.03) to V4 (0.16 AU ± 0.03), with p = 0.002 (Cohen’s d_z_ = −0.50). A detailed description of tonic pupillary responses is further provided in section 3.3.

**Figure 3.**
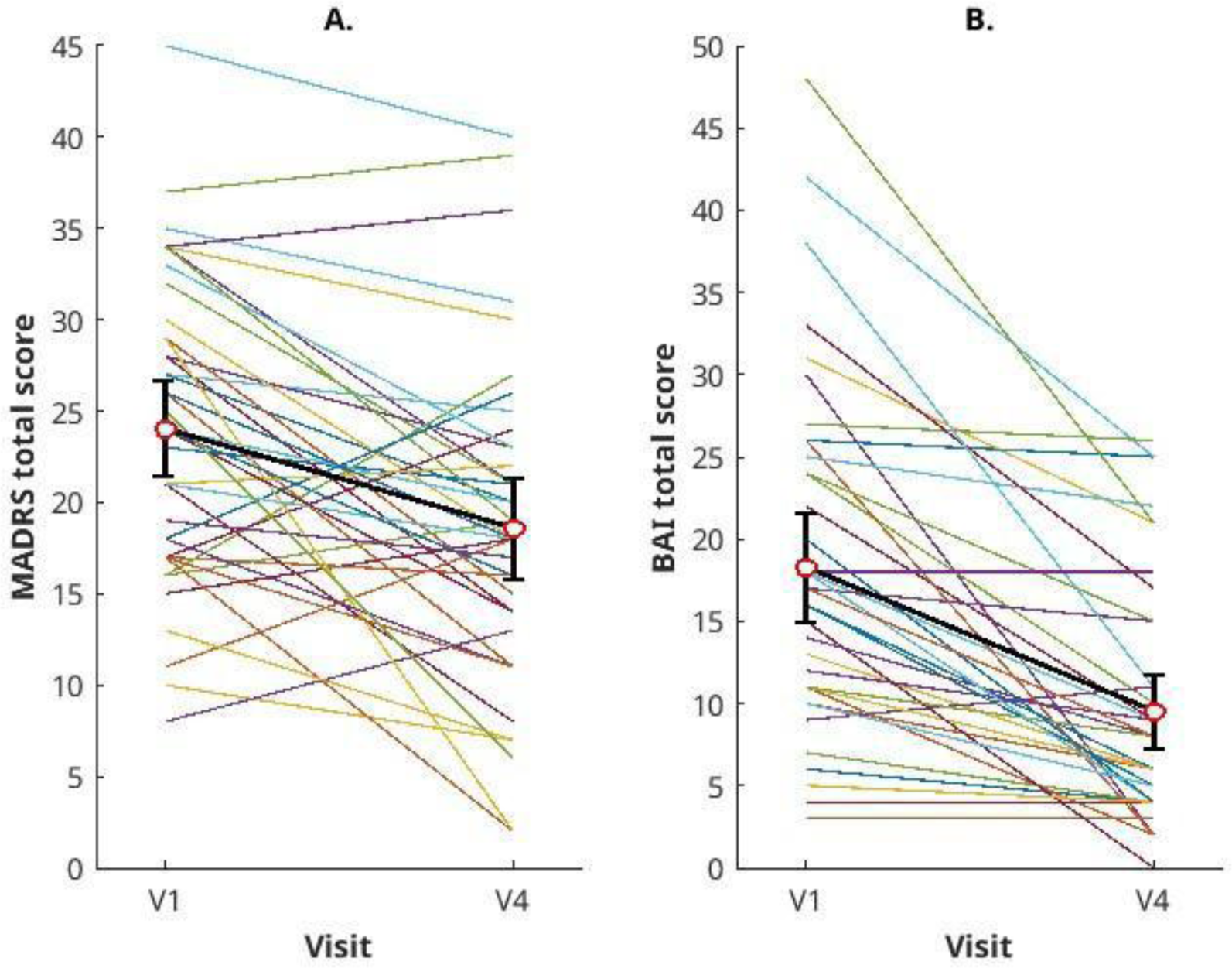
Individual trajectories for (A) Montgomery-Åsberg Depression Rating Scale (MADRS) and (B) Beck Anxiety Inventory (BAI) total scores from baseline (V1) to post-treatment (V4) visit. Each colored line represents an individual participant. Red circles and error bars represent the group mean and 95% confidence intervals at each visit. V1–V4 changes were assessed using two-sided paired-samples t-tests, with differences calculated as V4 minus V1.

**Figure 4.**
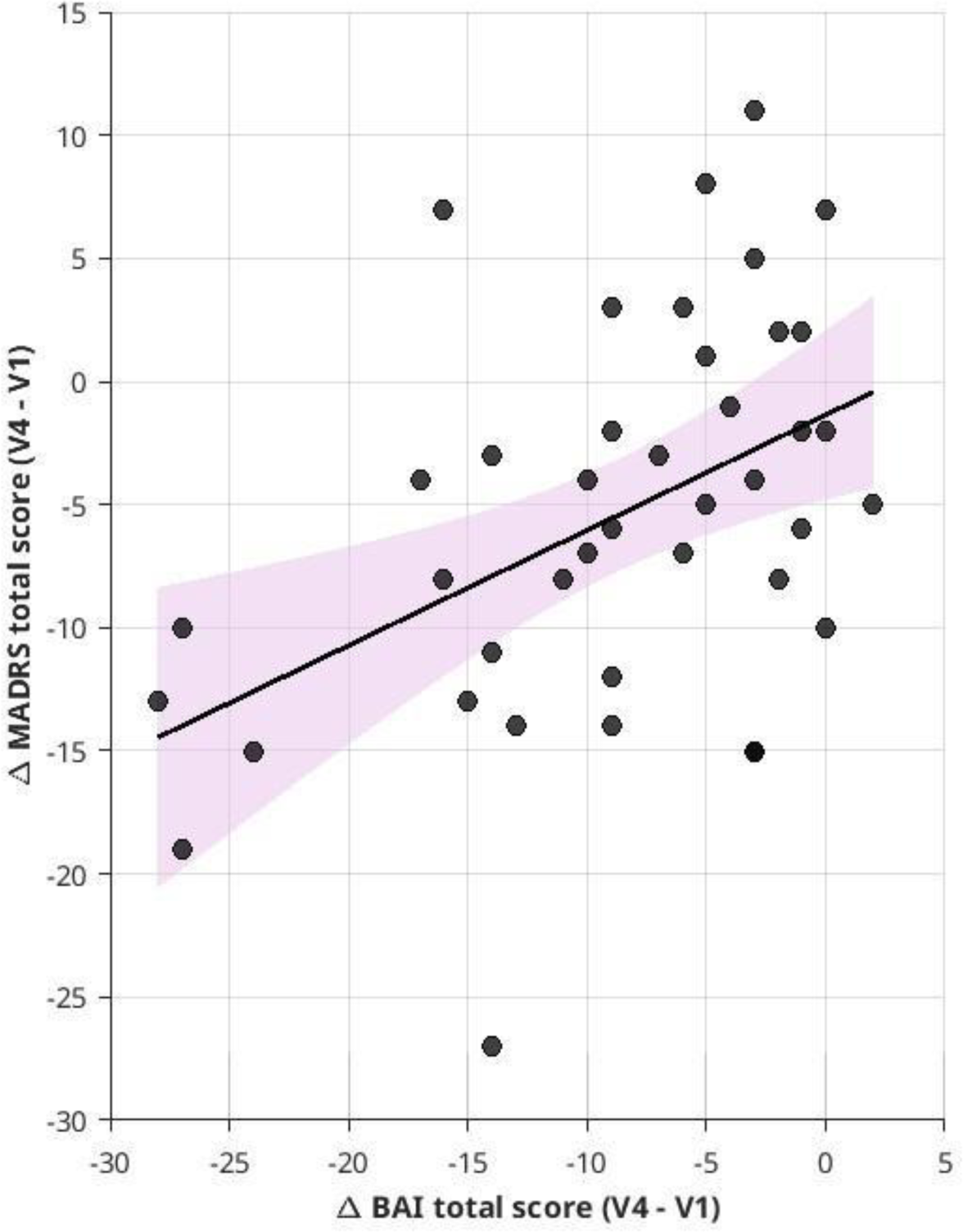
Association between the group difference Montgomery-Åsberg Depression Rating Scale (MADRS) and Beck Anxiety Inventory (BAI) total scores from baseline (V1) to post-treatment (V4) visit. Scatterplot showing the association between V1–V4 changes with each point representing one participant. The solid line represents the fitted linear regression line, and the shaded pink area indicates its 95% confidence interval. Greater reductions in depressive symptoms were associated with greater reductions in anxiety symptoms from V1 to V4 (Pearson’s r = 0.46, p = 0.002).

**Table 1.** Demographics and clinical data. Mean ± standard deviation values are reported for demographic and clinical parameters.

| Parallel arms | Active-sham<br>(n = 19) | Sham-active<br>(n = 22) | t/$\chi^2$, p-value, Cohen's effect size |
| --- | --- | --- | --- |
| Age (years) | 32.1 ± 12.07 | 32.86 ± 12.5 | t = -0.19, p = 0.84 |
| Age of onset (years) | 18.88 ± 7.5<sup>a</sup> | 20.19 ± 10.1<sup>b</sup> | t = -0.44, p = 0.65 |
| Number of episodes | 6.26 ± 9.6<sup>c</sup> | 3.75 ± 3.6<sup>d</sup> | t = 1.06, p = 0.29 |
| Psychotropic medication (load per visit) | V1 = 2 ± 1.9<br>V2 = 1.95 ± 1.9<br>V3 = 2.05 ± 1.9<br>V4 = 2.26 ± 2 | V1 = 2 ± 1.8<br>V2 = 2 ± 2<br>V3 = 1.91 ± 1.8<br>V4 = 1.95 ± 1.8 | Active–Sham: $\chi^2$ = 3.10, p = 0.37<br>Sham–Active: $\chi^2$ = 1.57, p = 0.66 |
| Sex | M = 7; F = 12 | M = 14; F = 8 | $\chi^2$ = 2.93, p = 0.08 |
| Education (years) | 16.57 ± 2.8 | 15.86 ± 2.1 | t = 0.91, p = 0.36 |
| MWTB total score | 28.84 ± 3.1 | 31 ± 2.3<sup>e</sup> | t = -2.42, p = 0.02 |
| **Whole sample (n = 41) clinical and physiological measures at V1 and V4** |  |  |  |
| Clinical and behavioral measures | **V1** | **V4** |  |
| MADRS total score | 24.07 ± 8.2 | 18.6 ± 8.8 | t = -4.37, p < 0.001, d<sub>z</sub> = -0.68 |
| BAI total score | 18.29 ± 10.3 | 9.53 ± 7.1 | t = -7.15, p < 0.001, d<sub>z</sub> = -1.12 |
| Physiological data | **V1** | **V4** |  |
| Mean pupil diameter (AU) | 0.18 AU ± 0.03 | 0.16 AU ± 0.03 | t = -3.25, p = 0.002, d<sub>z</sub> = -0.50 |

Medication load range for psychotropic administration: 0 = no intake/0mg; 1 = under reference range; 2 = in the reference range; 3 = above reference range.
Table legend: MDD = major depressive disorder; MWBT = Mehrfachwahl Wortschatz Intelligenztest; MADRS = Montgomery-Åsberg Depression Rating Scale; BAI = Beck Anxiety Inventory; AU = arbitrary units.
<sup>a</sup> n = 18
<sup>b</sup> n = 21
<sup>c</sup> n = 14
<sup>d</sup> n = 20
<sup>e</sup> n = 22

### 3.2 Linear mixed-effects analysis on pupil size-RSNs coupling

Across all seven networks considered for the pupil-RSNs coupling (visual, dorsal and ventral DMN, left and right ECN, SN, DAN+VAN system), the lECN was the only network showing a significant longitudinal change in its coupling with pupil size across visits (F(3,131) = 4.95, pFDR = 0.01, Figure 5). The longitudinal lECN pupil– brain coupling effect was robust to individual observations (Supplementary Materials, **Fig. S2**), remaining FDR-significant across all but one leave-one-observation-out analysis. Sensitivity analyses, including those based on standardized residuals, are described in the Supplementary Materials (**Fig. S2**). Additionally, the results of the LME analyses for all the other RSNs are seen in the Supplementary Materials (**Fig.S3**).

**Figure 5.**
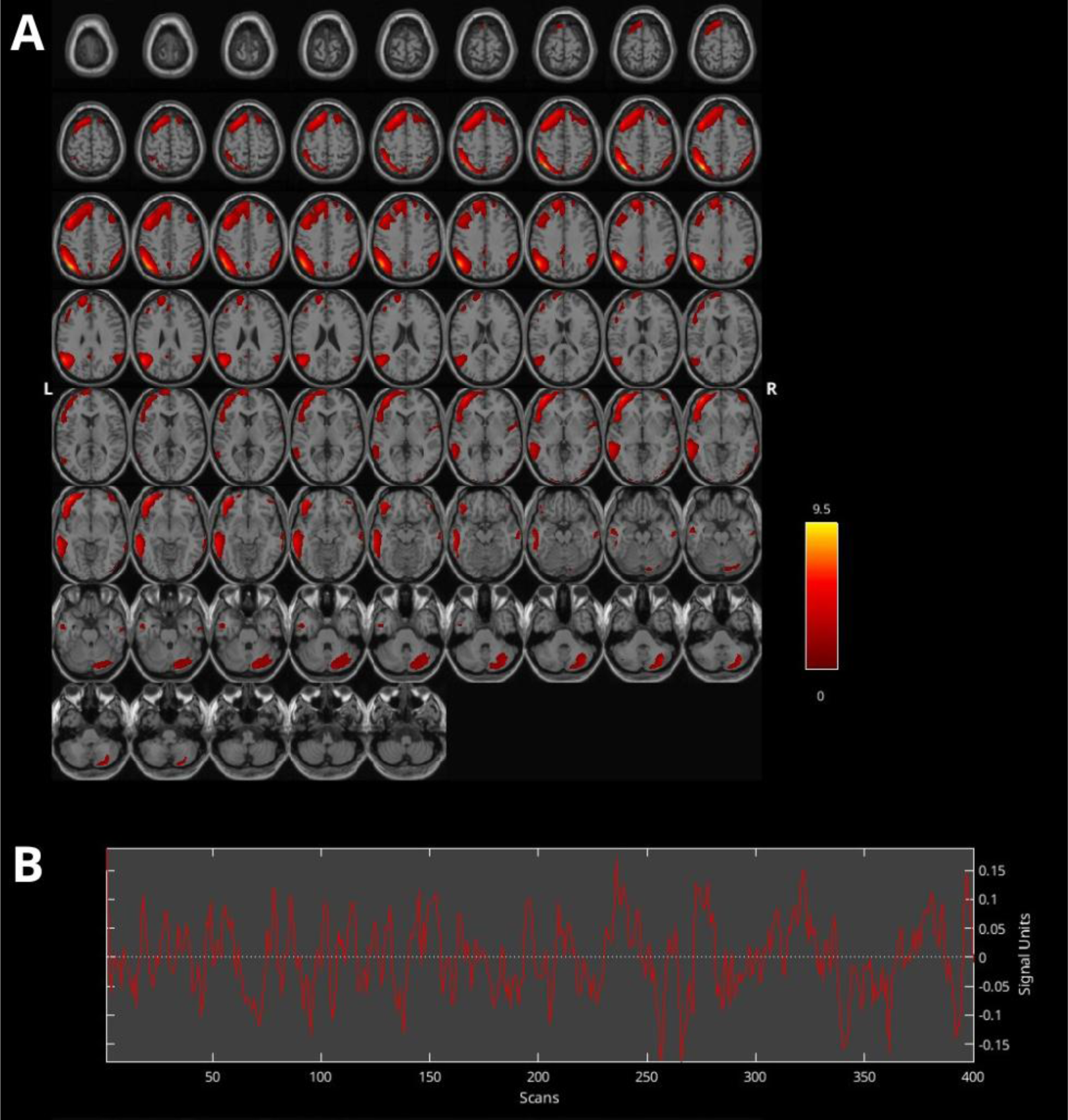
Left executive control network (lECN) z-scored statistical map (warm colors) derived from group independent component analysis (GIFT-ICA). The figure shows **(A)** the positive z-score map of the lECN independently of pupil size coupling, across all visits (V1, V2, V3, and V4) and all subjects, **(B)** with its corresponding averaged time course.

In **Figure 5**, for visualisation purposes and to highlight the spatial expression of the lECN, an axial positive z-scored map of the lECN derived from GIFT across all visits and all subjects, together with its corresponding averaged time course. In **Figure 6**, instead, we show significant longitudinal changes underscored by the coupling between pupil size and lECN across the whole treatment period.

**Figure 6.**
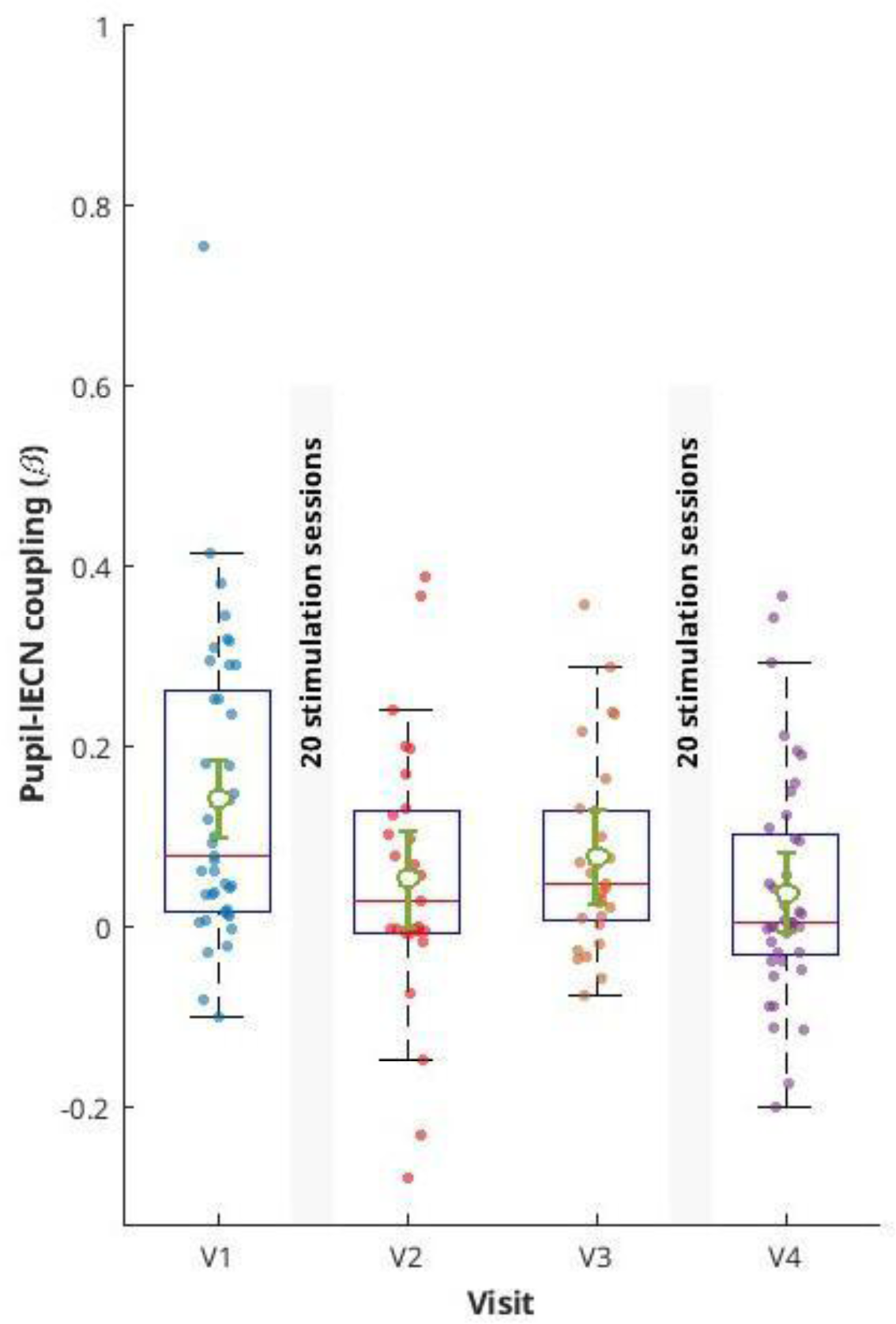
Linear mixed-effects model of pupil size-left executive control network (lECN) coupling across study visits. Box plots and jittered individual observations show the distribution of pupil–lECN coupling coefficients across study visits (V1, V2, V3, and V4). Open green circles with error bars represent model-estimated marginal means (EMMs) and 95% confidence intervals from the longitudinal linear mixed-effects model. The overall effect of Visit was assessed using the omnibus test of the Visit term. The analysis was adjusted for sequence, age, and sex as fixed effects, with subjects included as a random intercept. The analysis revealed a significant longitudinal effect of Visit on pupil size-lECN coupling (F(3,131) = 4.95, pFDR = 0.01). Each intervention week, comprising 20 stimulation sessions, is represented by a gray bar.

Exploratory analyses on this primary finding indicated that longitudinal changes in pupil size-lECN coupling did not differ significantly by intervention sequence (active-sham vs. sham-active), as reflected by the Visit x Sequence interaction (F(3,128) = 0.23, p = 0.86; **Fig.7**). The interaction remained non-significant across all leave-one-observation-out sensitivity analyses (p range = 0.74 - 0.98), and standardized residuals did not identify any observations with extreme residual values.

**Figure 7.**
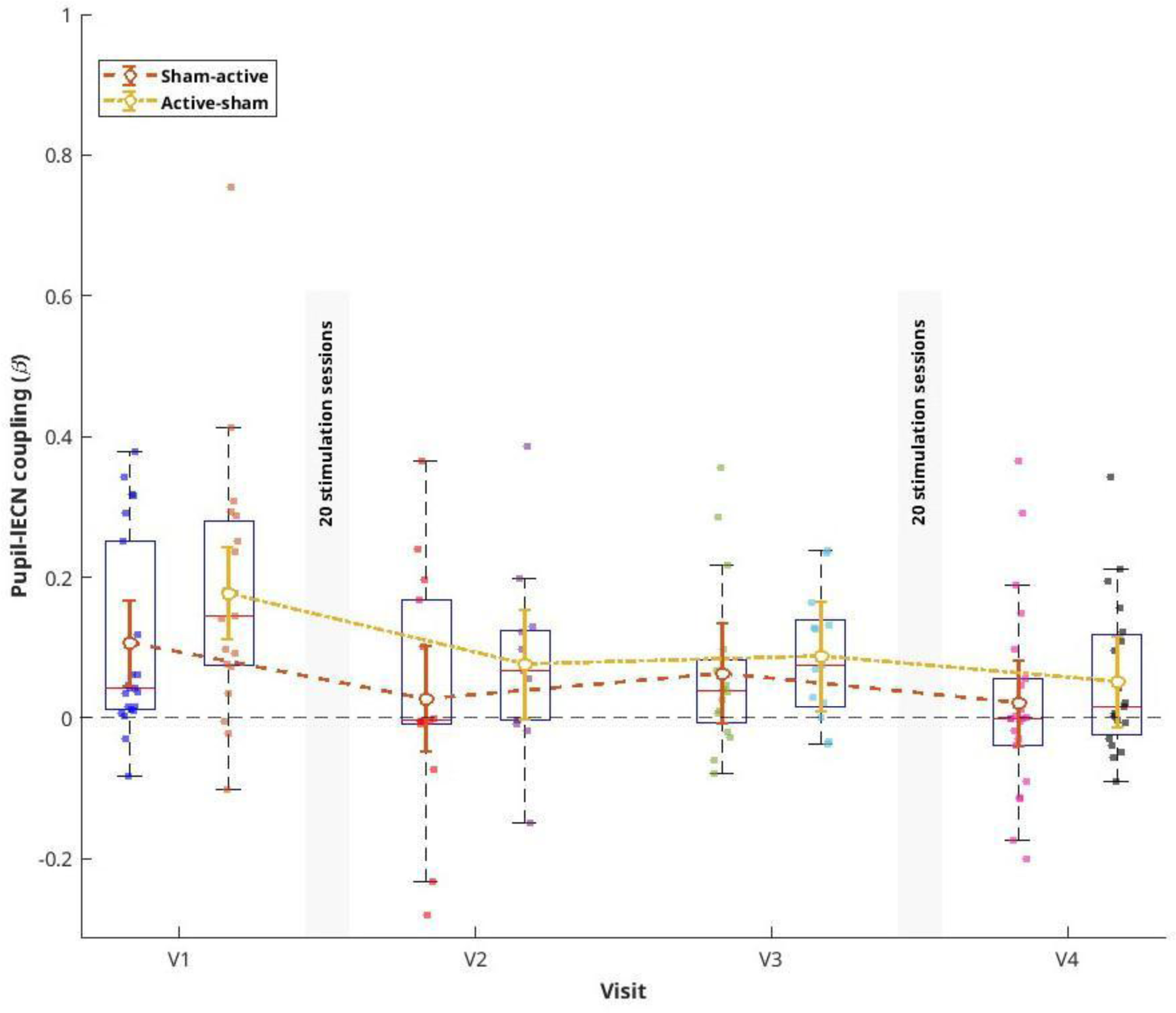
Exploratory linear mixed-effects model of pupil size-left executive control network (lECN) coupling and intervention order. Box plots and jittered individual observations show the distribution of pupil-left executive control network (lECN) coupling coefficients across visits (V1, V2, V3, and V4) for participants assigned to the sham-active and active-sham intervention sequences. Lines with open circles and 95% confidence intervals represent model-estimated marginal means (EMMs) from a linear mixed-effects model including Visit, Intervention sequence, the Visit × Sequence interaction, age, and sex as fixed effects, and subject as a random intercept. The horizontal dashed line indicates zero coupling. The Visit and the Visit × Sequence interaction was not statistically significant (F(3,128) = 0.23, p = 0.86), indicating no evidence that longitudinal changes in pupli-lECN coupling differ by intervention sequence. Each intervention week, comprising 20 stimulation sessions, is represented by a gray bar.

To further examine whether treatment response, defined by MADRS responder status (responders vs. non-responders), was associated with longitudinal changes in pupil–lECN coupling, we tested the Visit × Sequence × Responder status interaction. This interaction was significant (F(3,120) = 4.02, p = 0.009), indicating that the longitudinal pattern of pupil–lECN coupling differed as a function of treatment sequence and responder status. Significant lower-order interactions were also observed for Visit × Responder status (F(3,120) = 2.70, p = 0.04) and Sequence × Responder status (F(1,120) = 7.50, p = 0.007). Although one observation showed an elevated standardized residual, sensitivity analyses confirmed the robustness of the three-way interaction (p range = 0.003 - 0.03). Post-hoc analyses were therefore conducted to characterize the three-way interaction using Holm correction for the targeted follow-up comparisons. Among participants from sham-active intervention, the responder–non-responder difference showed its largest change relative to baseline at V2 (uncorrected p = 0.01), whereas among those from active-sham intevention, the corresponding difference was observed at V3 (uncorrected p = 0.02). However, neither comparison remained significant after Holm correction (p_Holm_ = 0.08 and 0.12, respectively). Thus, the significant omnibus interaction indicates the longitudinal pattern of pupil size-lECN coupling varied jointly by responder status and intervention sequence, whereas the specific visit- and sequence-level differences identified in the post-hoc analyses should be considered exploratory. These analyses are presented in more detail in the Supplementary Materials (**Fig. S4A– S4B**). In a separate exploratory analysis using MADRS as a continuous measure, changes in MADRS total scores from V1 to V4 were not significantly associated with corresponding changes in pupil size-lECN coupling (Pearson r = 0.14, p = 0.38). Leave-one-subject-out sensitivity analyses confirmed that the correlation between MADRS total scores and changes in pupil size-lECN coupling coefficients remained non-significant (p range = 0.22 - 0.57). This continuous-score analysis of changes between pre- and post-inteventions is presented in more detail in the Supplementary Materials (**Fig. S5**).

Finally, exploratory analyses of BAI total scores revealed that anxiety symptoms did not significantly moderate pupil size-lECN coupling in the full longitudinal LME (F(3, 127) = 0.34, p = 0.79). Furthermore, the Visit x Sequence x BAI interaction was not significant (F(3,120) = 0.44, p = 0.72), providing no evidence that intervention sequence further moderated the longitudinal association between anxiety symptoms and pupil-lECN coupling. Changes from V1 to V4 were also not significantly associated (Pearson r = 0.08, p = 0.60). For the full LME model, the Visit x BAI interaction remained non-significant in all leave-one-observation-out analyses (p range = 0.67 - 0.92). Similarly, the correlation between BAI total score and pupil size-lECN coupling coefficients remained non-significant in the sensitivity analyses (p = 0.80). Further details on the BAI analyses can be found in the Supplementary Materials (**Fig. S6** and **Fig. S7**).

### 3.3 Tonic pupil dynamics

Considering all four intervention-related visits, mean pupil diameter did not show a significant longitudinal change (F(3,131) = 2.16, p = 0.09). Inspection of standardized marginal residuals identified two observations with extreme values. Sensitivity analyses excluding these two observations resulted in a nominally significant longitudinal Visit effect (F(3,128) = 3.80, p = 0.01; Supplementary Materials **Fig. S8**). Notably, the LME analysis of tonic pupillary responses also revealed a significant lower-order effect of age (F(1,131) = 4.71, p = 0.03), which is further described in the Supplementary Materials (**Fig. S9**). Finally, longitudinal changes in mean pupil diameter were not significantly moderated by treatment sequence (F(3,128) = 0.51, p = 0.67; **Fig. S10**) or treatment response (F(3,127) = 0.84, p = 0.47; **Fig.S11A-S11B**). The Visit × Sequence × MADRS (responder status) was also not significant (F(3,120) = 1.95, p = 0.12). Sensitivity analyses yielded consistent results for the Visit × Sequence and Visit × MADRS (responder status) interactions (all p > 0.19), whereas the three-way interaction showed some sensitivity to the exclusion of individual observations (p range = 0.04 - 0.33). These moderation analyses are detailed in the Supplementary Materials (**Fig. S11A–S11B**). In a separate exploratory continuous-score analysis, changes in mean pupil diameter were not significantly associated with corresponding changes in MADRS total scores between the V1 and V4 time points (Pearson r = 0.09, p = 0.57; Supplementary Materials **Fig. S12**). This finding was robust to sensitivity analyses (p range = 0.25 - 0.87).

In contrast, a paired comparison of mean pupil diameter between V1 and V4 revealed a significant decrease (p = 0.002; **Table 1**; **Fig. 8**). No participant showed an extreme V1–V4 change score, and the result remained significant across all leave-one-subject-out sensitivity analyses (p range = 0.001 - 0.004).

**Figure 8.**
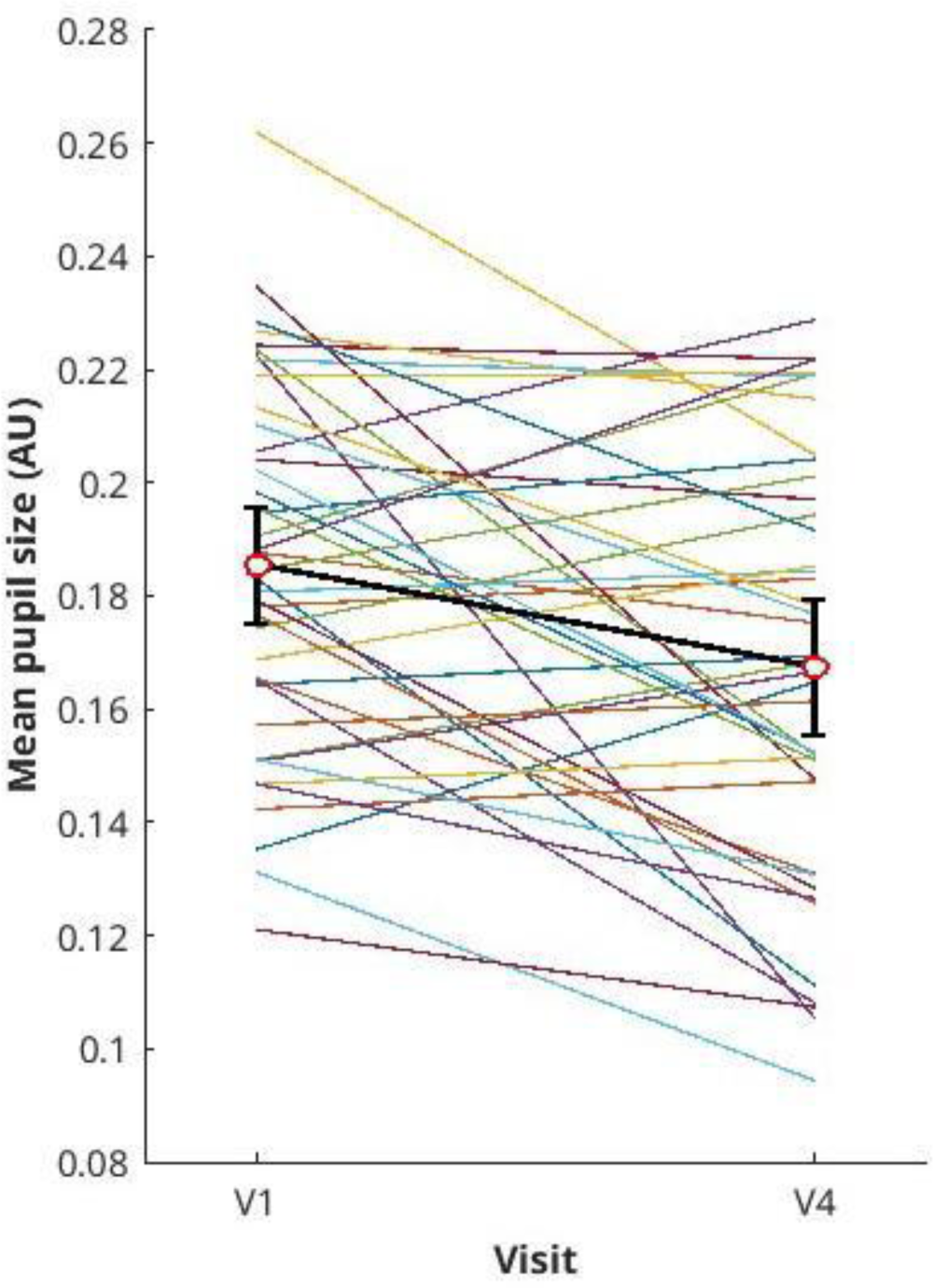
Longitudinal changes in mean pupil size from baseline (V1) to post-treatment (V4). Each colored line represents an individual participant’s within-subject trajectory from V1 to V4 for mean pupil diameter. Red circles and error bars represent the group mean and 95% confidence interval at each visit. V1–V4 changes were assessed using two-sided paired-samples t-tests, with differences calculated as V4 minus V1. Pupil diameter is expressed in arbitrary units (AU).

When additional exploratory analyses were conducted using BAI total scores, the full LME model did not reveal a significant Visit × BAI interaction in relation to longitudinal changes in mean pupil diameter across the four visits (F(3, 127) = 1.99, p = 0.11; see Supplementary Materials, **Fig. S13**). The three-way Visit x BAI score x Sequence interaction was also not significant (F(3, 120) = 0.47, p = 0.69). Sensitivity analyses showed that the Visit × BAI interaction was sensitive to the exclusion of individual observations, with one exclusion yielding p = 0.04 - consistent with the presence of an observation with elevated standardized residual. In contrast, the three-way interaction with intervention order remained non-significant across all individual-observation exclusions (p = 0.43 - 0.81).

Finally, Pearson correlation analyses of matched V1–V4 changes showed that greater improvement in anxiety symptoms was significantly associated with a greater decrease in mean pupil diameter (r = 0.45, p = 0.003; **Fig.9**). Neither change variable showed an extreme standardized change score, and the association remained significant across all leave-one-subject-out analyses (r = 0.33 - 0.51, p range = 0.001 - 0.03).

**Figure 9.**
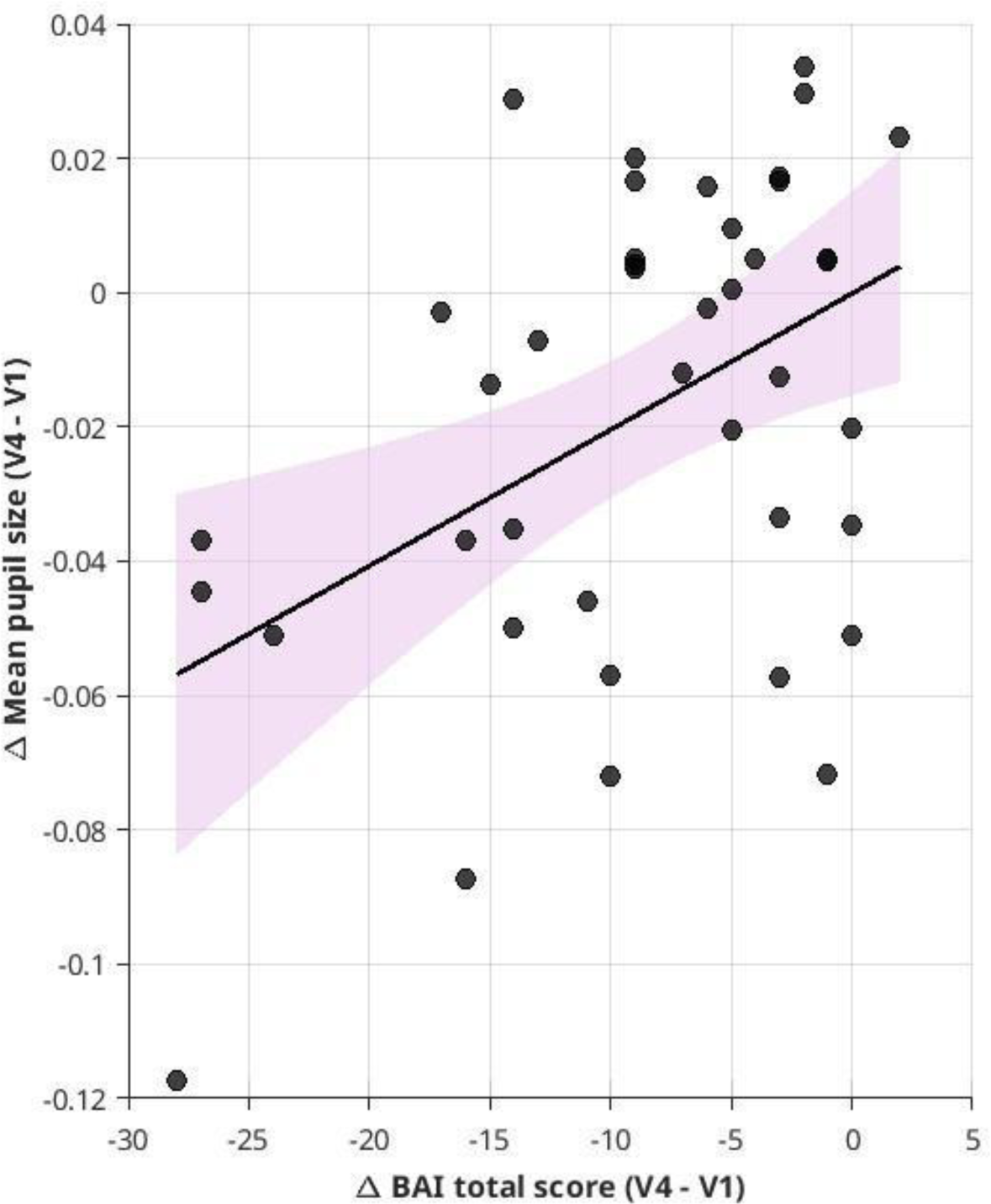
Association between changes in Beck Anxiety Inventory (BAI) scores and mean pupil size from baseline (V1) to post-treatment (V4) visit. Scatterplot showing the association between V4–V1 changes with each point representing one participant. The solid line represents the fitted linear regression line, and the shaded pink area indicates its 95% confidence interval. Greater improvement in anxiety symptoms was associated with a greater decrease in mean pupil diameter (Pearson’s r = 0.45, *p* = 0.003). Pupil diameter is expressed in arbitrary units (AU).

## 4 DISCUSSION

The current study aimed to provide insights into whether tonic pupil dynamics may serve as a potential peripheral marker of longitudinal changes in their coupling with resting state functional networks during iTBS treatment in MDD.

Among the RSNs considered, lECN–pupil coupling was the only brain–body association showing a significant longitudinal change across the four study visits. Specifically, we observed an overall decrease in the positive tonic pupil-related coupling with the lECN from baseline (pre-treatment) to the end of treatment, across the overall treatment period. Of note, the absence of significant longitudinal effects for the other RSNs should not be interpreted as an absence of pupil–network coupling: rather, these networks showed varying positive or negative coupling with the pupil signal, without evidence for a systematic change in coupling across visits. Thus, among the networks examined, the longitudinal change in pupil-RSN coupling was specific to the lECN, rather than reflecting a generalized change across RSNs. Given the spatial organisation of the lECN as a cognitive control network spanning frontal and parietal regions, including the left dlPFC - the cortical target of stimulation-, and the lateralisation of the network, this primary finding may be particularly relevant to the intervention. Specifically, the ECN is involved in higher-order cognitive processes spanning from WM, cognitive control reserves, and executive functions, including decision-making mechanisms (Heine et al., 2012; Menon & Uddin, 2010; Schneider et al., 2016). Of note, in the context of the so-called cognitive model of depression, studies have evidenced the role of the ECN in orchestrating cognitive control as well as externally directed attention or internal mentation in MDD: under this perspective, the ECN has been implicated in cognitive control deficits and altered attention related processes frequently observed in the disorder, including difficulties in actively recruiting cognitive control resources, stronger self-referential thoughts, and impaired goal-directed control of attention toward external stimuli (Dichter et al., 2009; Everaert et al., 2012; Kaiser et al., 2015; Ochsner et al., 2004; Prompiengchai & Dunlop, 2025).

Pupil fluctuations have been shown to covary with activity across several large-scale RSNs, including networks involved in arousal and cognitive control processes. For example, studies in healthy individuals have evidenced that pupil diameter is actively coupled with frontoparietal regions that functionally overlap with the ECN, endorsing cognitive load and attentional focus allocation in task-related contexts (Alnæs et al., 2014; Aston-Jones & Cohen, 2005; Schneider et al., 2016; Siegle, Steinhauer, Stenger, et al., 2003). At the level of large-scale network organisation, orienting and alerting processes have been linked to neural systems involved in pupillary control, including the SC and LC–NE systems. Within this broader framework, the ECN, together with its connections to sensory regions, may then contribute to the top-down control of attention and thereby influence tonic and transient changes in pupil diameter (Strauch et al., 2022).

In depression, previous studies have suggested that pupillary measures may provide a useful psychophysiological marker of neurocognitive alterations, particularly those related to executive functioning and cognitive control (Fietz et al., 2024; Jones et al., 2015; Siegle et al., 2004). However, to the best of our knowledge, among intervention studies, including those involving rTMS, relatively little attention has been given to the interplay between pupil dynamics and RSNs in MDD (Citrenbaum et al., 2023; Fietz et al., 2024; Jones et al., 2015; M. K. Leuchter et al., 2026; Noyes et al., 2026; Siegle et al., 2004). Thus, the present findings might be interpreted as evidence of an initially stronger positive coupling between tonic pupillary responses and lECN in MDD: one possible interpretation is that the altered pupil size-lECN coupling observed over the treatment period may reflect changes in the dynamic relationship between cognitive-control processes and the regulation of arousal-related pupillary fluctuations at rest (Aminihajibashi et al., 2019; Jones et al., 2015). Of note, the overall changes characterizing the sustained reduction in the positive interplay between the lECN and tonic pupillary responses may be informative, given that cognitive deficits and persistent negative self-referential or ruminative processes are core features of MDD and reflect difficulties in flexibly integrating externally directed information with internally focused processing (Disner et al., 2011; Schiena et al., 2021; Schwert et al., 2017). However, this interpretation remains speculative, as the present study did not directly assess the dynamic integration of external information or the cognitive mechanisms underlying pupil size-lECN coupling. Whether the longitudinal change in pupil size-lECN coupling reflects an adaptive change in the coordination between cognitive-control and arousal-related processes remains still unclear (Aminihajibashi et al., 2019; C. L. Jones et al., 2015). Due to the limited literature on ECN–pupil coupling in depression at rest, particularly in the context of neuromodulatory interventions such as iTBS, future studies should examine whether longitudinal changes in lECN–pupil coupling are related to the response to treatment and whether they reflect an adaptive change. Anxiety should also be targeted as a potentially mediating factor in this relationship, and a healthy control group should be included for comparison.

Moreover, the physiological findings of the current study may provide further context for interpreting the longitudinal change in lECN–pupil coupling. We did not find systematic longitudinal changes of mean pupil diameter when considering the full four-visit treatment period. Under this perspective, we observed a significant association between age and pupil size, consistent with the established influence of demographic factors on pupillary measures in rTMS intervention studies, as well as with age-related influences on pupillary measures and previously reported changes in adrenergic/noradrenergic function in depression (e.g., Leuchter et al., 2026; Sokolski et al., 2000). Nevertheless, it is worth noting that when restricting the analysis to the time-matched comparison between baseline and the end of treatment, mean pupil diameter showed a significant decrease. Accordingly, while task-based studies have reported sustained or enhanced pupil dilation due to the occurrence of emotionally and cognitively demanding processing mechanisms, few studies conducted during resting conditions have suggested that depressed patients may show larger baseline pupil diameter, especially when compared with age-matched healthy individuals (Schumann et al., 2017; Siegle, Steinhauer, Carter, et al., 2003; Sokolski et al., 2000). In particular, larger pupil diameter has been proposed to reflect an overall increased demand on the noradrenergic system often observed in depression, underscoring the role of LC-NE circuit in the regulation of sympathetic and parasympathetic outflow in patients (Ryabinin et al., 2013; Samuels & Szabadi, 2008; Schumann et al., 2017; Sokolski et al., 2000). To the best of our knowledge, within the context of rTMS, relatively few studies have examined pupillary measures as potential physiological markers of treatment-related change in depression (Citrenbaum et al., 2023; M. Leuchter et al., 2025; M. K. Leuchter et al., 2026). Available evidence has linked treatment outcome to alterations in autonomic and pupillary measures, including reductions in resting pupil diameter following rTMS and greater pupil constriction responses, with greater pupillary constriction and related adaptive autonomic changes being associated (Citrenbaum et al., 2023; M. Leuchter et al., 2025; M. K. Leuchter et al., 2026). Thus, the reduction in resting pupil diameter observed between baseline and post-treatment visits may be consistent with previously reported treatment-related changes in pupillary and autonomic measures. However, the present study focused on mean pupil diameter at rest and did not assess more specific indices of pupillary reactivity, such as task-evoked constriction or dilation responses, nor did it include a healthy control group with which to directly assess depression-related differences in tonic pupil size. Thus, future studies should combine resting-state and task-evoked pupillometry with complementary measures of autonomic and noradrenergic function to determine whether longitudinal changes in tonic pupil size during rTMS reflect broader alterations in arousal and autonomic regulation across the treatment period (M. Leuchter et al., 2025; M. K. Leuchter et al., 2026).

Furthermore, another context for interpreting this physiological endpoint change may be provided by the relationship between tonic pupillary and anxiety levels, a clinical dimension often present in patients with depression (Aminihajibashi et al., 2019; Jones et al., 2015). We therefore additionally examined whether changes in anxiety symptoms - as measured with BAI total scores - were associated with changes in pupil diameter as well as in the pupil size-lECN coupling. Accordingly, we found that greater reductions in BAI scores were linked to greater reduction in mean pupil diameter between baseline and the end of treatment. This individual-difference association suggests that tonic pupillary dynamics in MDD may vary in relation to anxiety related states: for example, other studies in the field have proposed that a reduction in pupil diameter might be positively associated with a reduction of anxiety symptoms in depressive patients (Aminihajibashi et al., 2019; FERENCOVA et al., 2021; Garner, 2026; Hu et al., 2016; Jones et al., 2015). However, it is worth noticing that in the current study changes in pupil diameter were not significantly associated with changes in depressive symptom severity, or in pupil size-lECN coupling. Nonetheless, reductions in anxiety symptoms alone were themselves associated with reductions in depressive symptoms, as measured at MADRS: this individual-difference association might further suggest that tonic pupillary dynamics may vary with anxiety-related emotional state during treatment. One possible explanation is that tonic pupil size is more closely related to anxiety-related (hyper)arousal and defensive processes, including the sympathetic “fight-or-flight” response, than to depressive symptom severity per se (Aston-Jones & Cohen, 2005; Atzori et al., 2016; Joshi et al., 2016). Overall, these findings highlight the potential relevance of considering changes in anxiety states when investigating longitudinal alterations in pupillary dynamics during depression (Aminihajibashi et al., 2019; Jones et al., 2015). However, this association was based on individual changes between only two pre- and post-treatment time points: thus, future studies should examine more directly whether changes in anxiety and other affective states are associated with longitudinal alterations in tonic pupillary dynamics across the treatment period.

Finally, exploratory analyses of treatment order indicated that pupil size-lECN finding was not dependent on treatment sequence. Specifically, the longitudinal change in pupil size-lECN coupling remained evident irrespective of whether participants received active or sham stimulation first. Furthermore, no effects of treatment order were observed in the tonic modulation of mean pupil diameter, independently of its coupling with lECN. Few studies in healthy individuals have evidenced differences between active vs. sham conditions in pupillary responses, as outlined by larger pupillary responses in active condition, and absent effects in sham; however, to the best of our knowledge, the role of treatment order is not widely investigated in the context of rTMS-pupillary responses in depression (Citrenbaum et al., 2023; M. Leuchter et al., 2025; M. K. Leuchter et al., 2026; Niehaus et al., 2001; Phylactou et al., 2026; Weijs et al., 2025). It is also important to note that the absence of treatment order effects could be related to differences in rTMS protocols, which could limit the generalizability of the findings to the current study design (Phylactou et al., 2026; Weijs et al., 2025). Thus, future studies should further examine whether treatment sequence is associated with longitudinal changes in pupil dynamics and their coupling with major RSNs.

## 5 LIMITATION AND FUTURE DIRECTIONS

Several caveats should be considered when interpreting the present findings. First, the relatively small sample size and the unbalanced number of observations across study visits may have reduced statistical power, particularly for the exploratory analyses, thereby potentially reducing the generalisability of the findings. Although linear mixed-effects models can accommodate unequal numbers of observations across participants, the reduced number of observations at individual visits may nevertheless have limited the precision with which longitudinal and interaction effects could be estimated.

Second, given the exploratory nature of the present study, we focused primarily on tonic pupillary dynamics as indexed by mean pupil diameter. Consequently, we did not comprehensively consider other dimensions of pupillary responses, including phasic measures or more specific indices of pupillary reactivity. These complementary measures may provide additional information on autonomic regulation and its relationship with large-scale resting-state networks, as well as on temporal changes in pupillary responses that may complement the observed longitudinal changes in pupil–RSN coupling in MDD (Citrenbaum et al., 2023; M. K. Leuchter et al., 2026; Schneider et al., 2016; Yellin et al., 2015).

Third, we used an ICA-based approach focused on predefined RSNs of interest, selected based on previous evidence concerning large-scale functional networks in MDD and the relationship between pupillary dynamics and resting-state network activity. While this approach allowed us to test a priori network-level hypotheses, it does not capture other forms of functional connectivity or network dynamics that could be identified using complementary approaches, such as seed-based, dynamic, or effective-connectivity analyses (Jensen et al., 2024; Prompiengchai & Dunlop, 2025; Razi et al., 2017; Schiena et al., 2021; Schneider et al., 2016; S. Wang et al., 2026; Yellin et al., 2015).

Fourth, the primary finding encompassing pupil size-lECN coupling was identified as an overall longitudinal effect across the whole study period. Although, to the best of our knowledge, this is one of the first studies to investigate the interplay between pupillary size and major RSNs in MDD using this methodological approach, future studies should further test whether treatment order (active vs sham) as well as the influence of other anamnestic factors (e.g., age) play a potential role in modulating pupil-related resting state functional networks associations during treatment (M. K. Leuchter et al., 2026; Phylactou et al., 2026).

Finally, the absence of a healthy control group and the acquisition of pupillometry exclusively during resting-state conditions limited our ability to determine whether the observed longitudinal changes were specific to MDD and to relate resting-state pupil–RSN coupling to affective processes. Future studies combining patients and healthy controls could provide further insight into longitudinal changes in pupil–RSN coupling (Aminihajibashi et al., 2019; Fietz et al., 2024; Jones et al., 2015; Siegle et al., 2004).

## 6 CONCLUSIONS

In conclusion, the present study provides evidence that tonic pupillary dynamics are coupled with IECN activity - which notably includes the left dlPFC as cortical target of stimulation - in MDD and that this coupling is reduced during the intervention. Given the role of the IECN in higher-order cognitive control and its relationship with arousal-related pupillary dynamics, this finding suggests that the link between tonic pupil fluctuations and top-down cognitive control over arousal-related mechanisms underlying tonic pupillary responses might change over time, irrespective of treatment condition. Furthermore, the degree of reduction in pupil diameter alone is found to be related to a reduction in anxiety symptoms. This could provide further insights into how disorder-defining affective traits could potentially moderate tonic pupillary dynamics in MDD, considering also the positive association between reduction of anxiety symptoms and alleviation of depressive symptoms between pre and post-treatment (Aminihajibashi et al., 2019; Citrenbaum et al., 2023; N. P. Jones et al., 2015; M. K. Leuchter et al., 2026; Schiena et al., 2021; Schneider et al., 2016; Yellin et al., 2015).

## Data Availability

All data produced in the present study are available upon reasonable request to the authors

## Acknowledgements

We would like to thank the support from the German Federal Ministry of Education and Research (Bundesministerium für Bildung und Forschung, BMBF: 01 ZX 1507, “PreNeSt - e:Med”) and the Department of Psychiatry and Psychotherapy of Jena University Hospital.

## CRediT authorship contribution statement

**Monica Di Giuliano**: Conceptualization, Methodology, Investigation, Data Curation, Writing - Original Draft, Writing - Review and Editing, Visualization. **Andy Schumann**: Methodology, Data Curation, Writing-Review and Editing. **Martin Walter:** Writing - Review and Editing. **Schmidt-Samoa Carsten**: Data Curation, Writing - Review & Editing. **Roberto Goya-Maldonado**: Conceptualization, Methodology, Investigation, Data Curation, Supervision, Writing - Review and Editing, Project administration.

## Funding

This work was funded by the German Federal Ministry of Education and Research (Bundesministerium für Bildung und Forschung, BMBF: 01 ZX 1507, “PreNeSt - e:Med”) and the Department of Psychiatry and Psychotherapy of Jena University Hospital.

## Declaration of Competing Interest

The authors declared that they do not have conflicts of interest.

## Notes

### Competing Interest Statement

The authors have declared no competing interest.

### Author Declarations

The study protocol complied with the Declaration of Helsinki and was approved by the Ethics Committee of the University Medical Center Goettingen (UMG).

