## Supplementary Materials for "Pupil size-resting state network coupling in major depressive disorder: a longitudinal iTBS study"

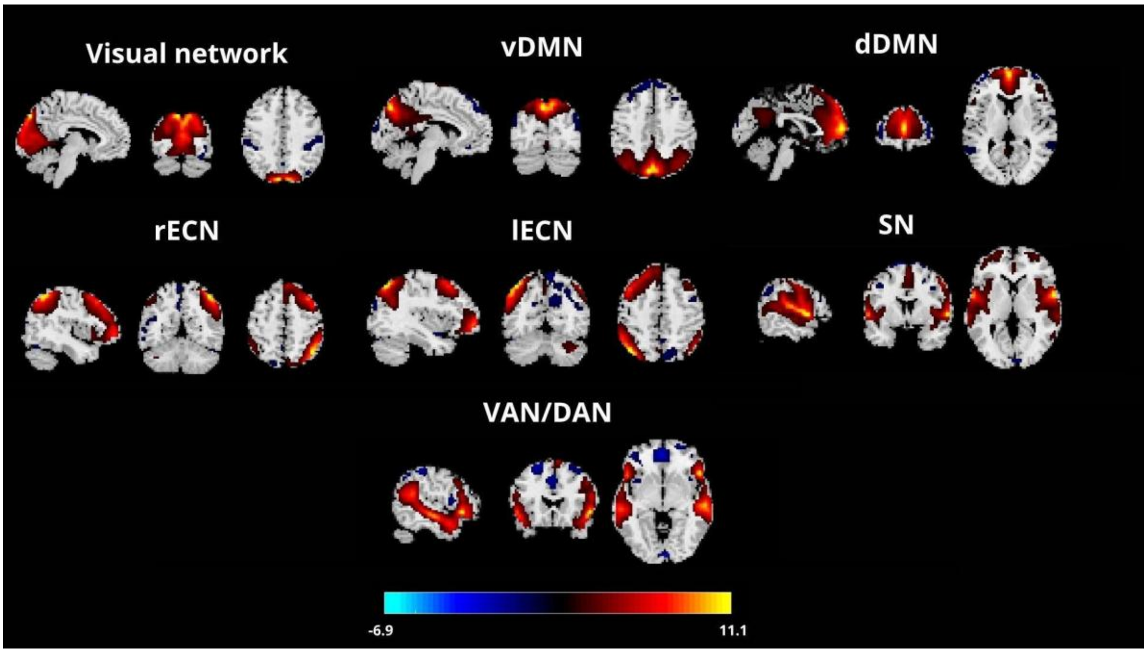


**Figure S1 Seven canonical resting state networks of interest, selected for group-level analyses from spatial sorting with Group Independent Component Analysis (GIFT-ICA).** The 20 independent components (ICs) yielded from GIFT-ICA on resting state data were spatially sorted using a canonical resting state network (NeuroMark 2.2 template) and then filtered for brain networks of interest, after visual inspection. The networks selected were: visual network; ventral default mode network (vDMN); dorsal default mode network (dDMN); right and left executive control network (rECN/lECN); salience network (SN), and finally attentional network in its dorsal and ventral parcellations (DAN+VAN). The figure shows the z-shifted maps in sagittal, coronal and axial view of the seven selected components, across all visits and all subjects (V1 V2 V3 V4).

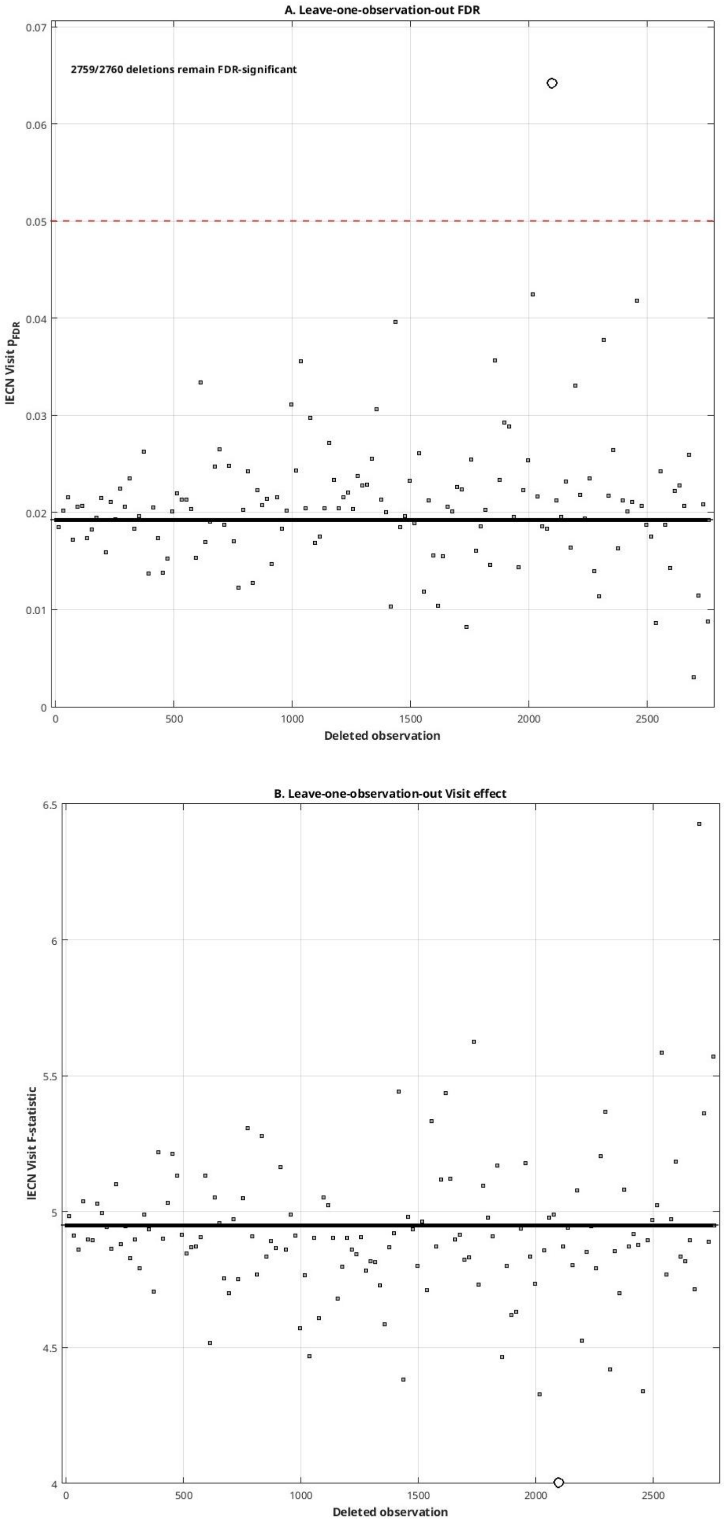


**Figure S2 Sensitivity of the longitudinal left executive control network (lECN) pupil size–brain coupling effect to individual observations. (A)** FDR-adjusted p-values for the longitudinal Visit effect in left executive control network (lECN) obtained after sequentially removing each of the 2,760 component-observations from the full dataset. The horizontal dashed line indicates the significance threshold of pFDR = .05, and the solid line indicates the FDR-adjusted p-value from the primary analysis (pFDR = 0.01). **(B)** Corresponding F-statistics for the Visit effect in the lECN. The solid line indicates the F-statistic from the primary analysis (F(3,131) = 4.95). The highlighted observation corresponds to one observation at baseline (V1); its removal resulted in pFDR = 0.06, whereas the effect remained FDR-significant after removal of the other 2,759 observations.

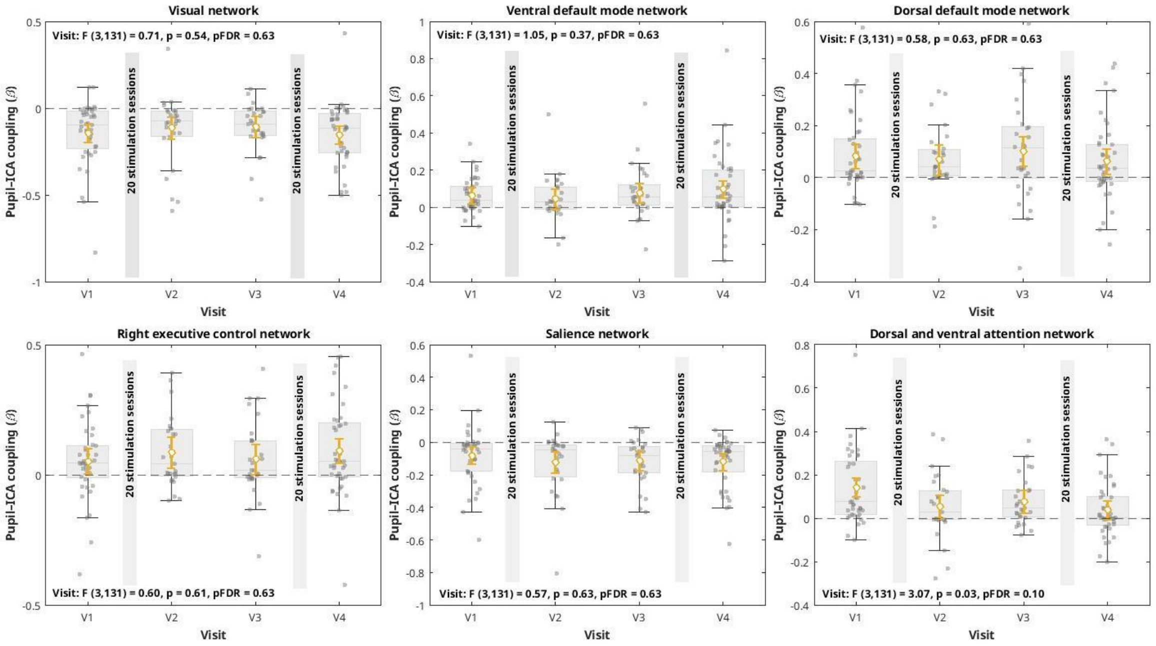


**Figure S3 Linear mixed-effects model on pupil size-resting state networks coupling across study visits.** Boxplots and jittered individual observations show the distribution of pupil–resting state networks coupling coefficients across study visits (V1 V2 V3 V4). Open yellow circles with error bars represent model-estimated marginal means (EMMs) and 95% confidence intervals from the longitudinal linear mixed-effects model. The overall effect of Visit was assessed using the omnibus test of the Visit term: the analysis is adjusted for sequence, age, and sex as fixed effects, with subjects included as a random intercept. F statistics and corresponding FDR corrected p-values for each pupil-resting state network (Independent Component Analysis - ICA) are reported, accordingly. Each intervention week, comprising 20 stimulation sessions, is represented by a gray bar.

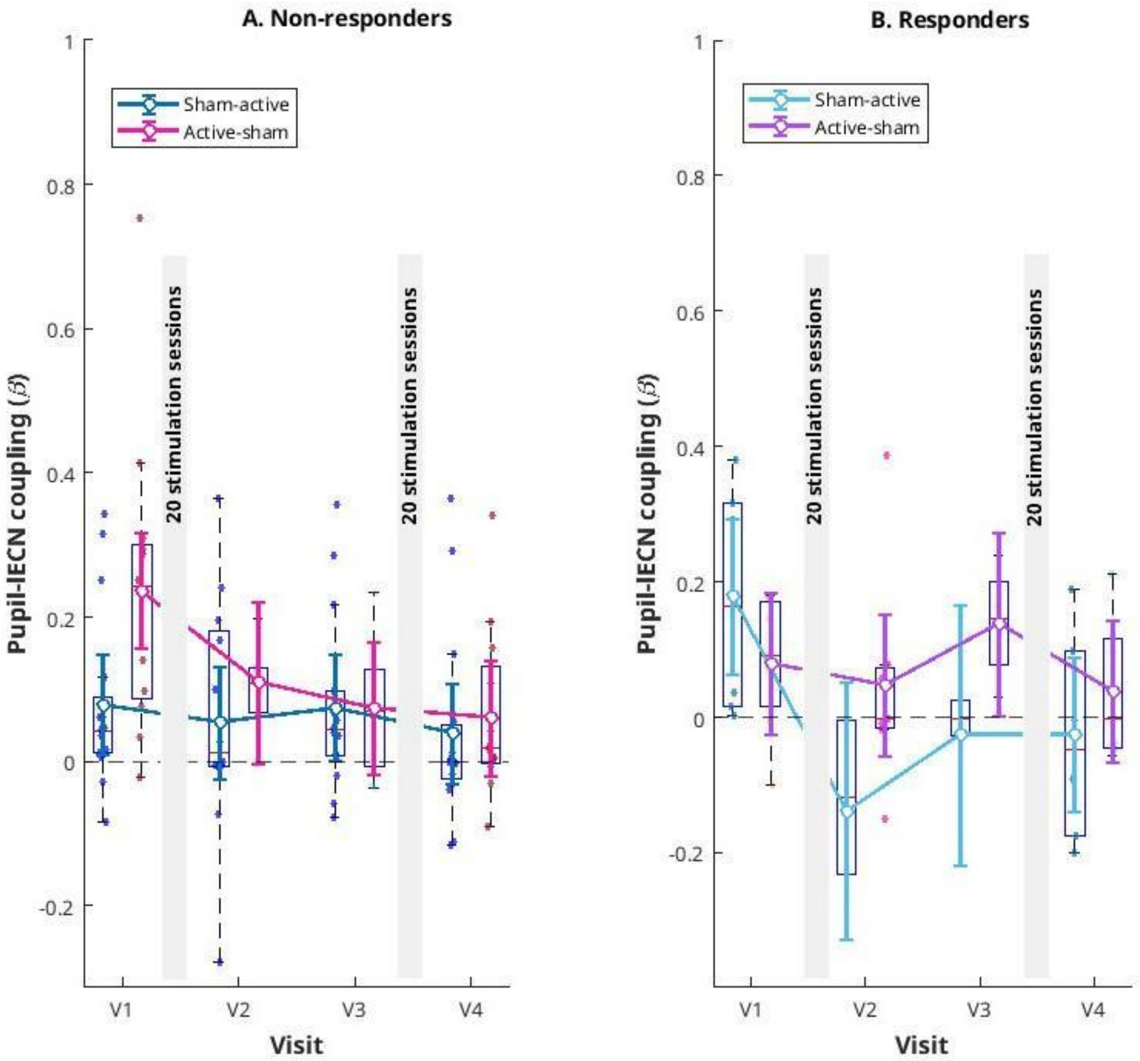


**Figure S4 Explorative linear mixed-effects model on pupil size-left executive control network (lECN) coupling and Montgomery-Åsberg Depression Rating Scale-MADRS x treatment order interaction effect.** Panels show pupil size - left executive control network (lECN) coupling across study visits, for non-responders (**panel A**) and responders (**panel B**). Boxplots and jittered individual observations represent the raw data. Lines with open circles and 95% confidence intervals represent model-estimated marginal means (EMMs) for the sham-active and active-sham treatment sequences. The longitudinal effects of Visit and treatment sequence were evaluated within the linear mixed-effects model, with the Visit × Sequence × Montgomery-Åsberg Depression Rating Scale (MADRS) interaction assessing whether longitudinal sequence effects differed according to response status. The horizontal dashed line indicates zero coupling. The analysis is adjusted for age and sex as fixed effects, with subjects included as a random intercept. Overall, we show a significant exploratory omnibus interaction indicating a difference in the longitudinal pattern across responder status and treatment sequence ($F(3,120) = 4.02$, $p = 0.009$). Each intervention week, comprising 20 stimulation sessions, is represented by a gray bar.

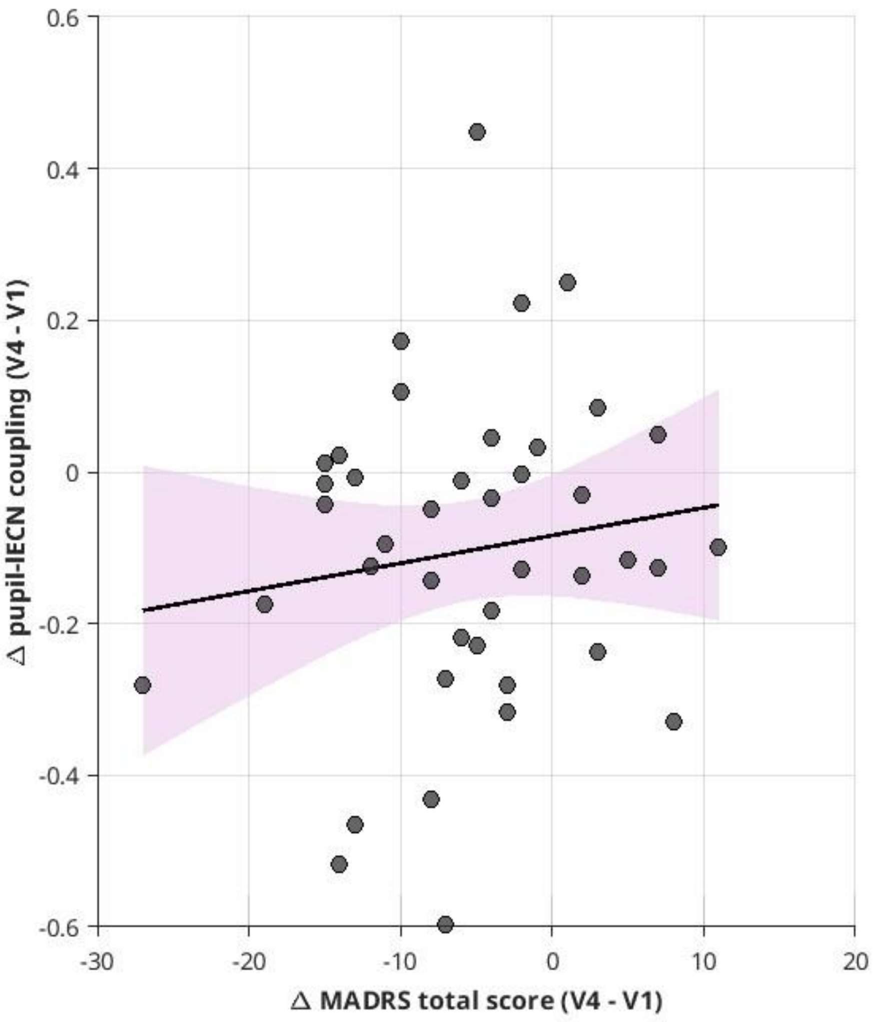


**Figure S5 Association between changes in depressive symptoms (Montgomery–Åsberg Depression Rating Scale - MADRS) and left executive control network (lECN) pupil coupling.** Scatterplot showing the association between changes in Montgomery–Åsberg Depression Rating Scale (MADRS) scores and changes in pupil size - left executive control network (lECN) coupling from baseline (V1) to the post-crossover visit (V4). Each point represents one participant. The solid line represents the fitted linear regression line, and the shaded pink area indicates its 95% confidence interval. Changes in MADRS scores were not significantly associated with changes in pupil–lECN coupling (Pearson's $r = 0.14$, $p = 0.38$).

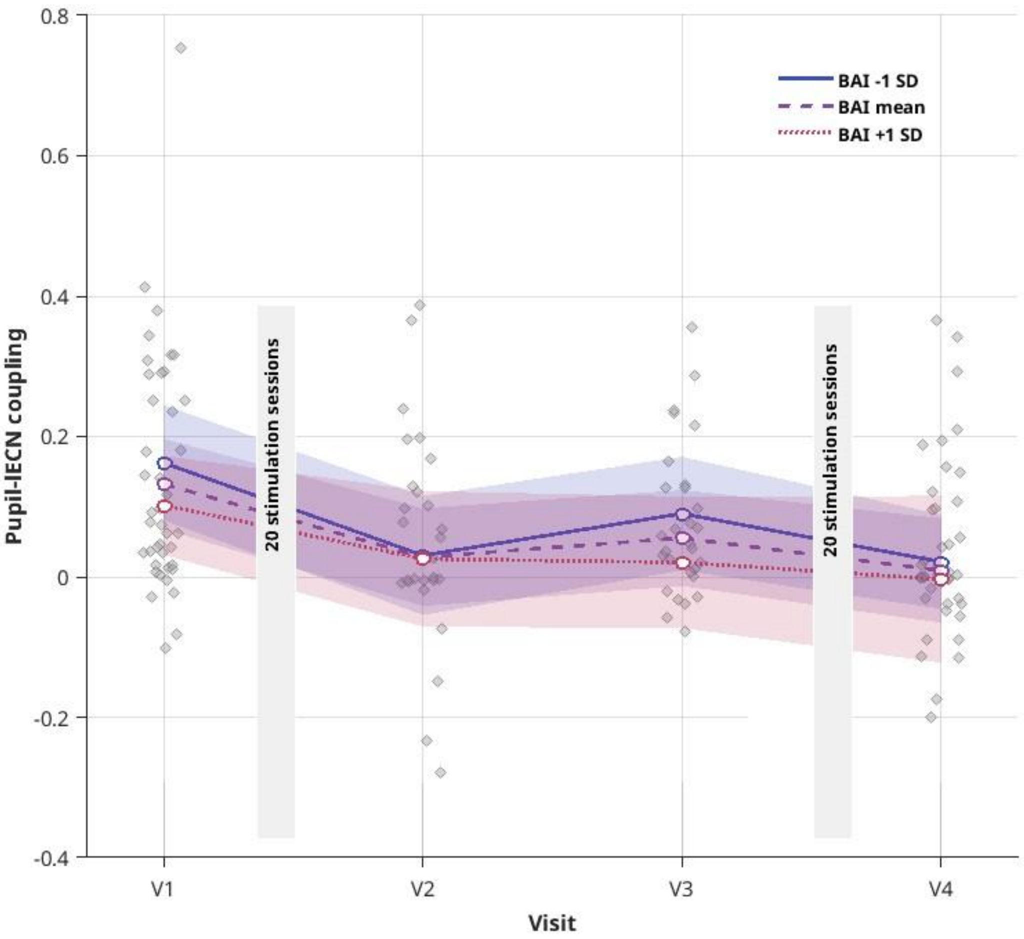


**Figure S6 Longitudinal pupil–left executive control network coupling as a function of Beck Anxiety Inventory (BAI) symptom level.** Observed pupil–left executive control network (lECN) coupling values are shown as individual points at each visit, with jitter applied horizontally for visualization. Colored lines and points represent estimated marginal means (EMMs) from the longitudinal linear mixed-effects model at the mean Beck Anxiety Inventory (BAI) score and at ±1 SD from the mean BAI total score; shaded areas indicate the corresponding 95% confidence intervals. The EMMs represent model-based predictions for representative levels of the continuous BAI moderator. The model included Visit × BAI interaction, treatment sequence, age, and sex as fixed effects, with a subject-level random intercept. Left-ECN-pupil coupling was shown to be not significantly moderated by changes in BAI total scores, across all visits ($F(3,127) = 0.34$, $p = 0.79$). Each intervention week, comprising 20 stimulation sessions, is represented by a gray bar.

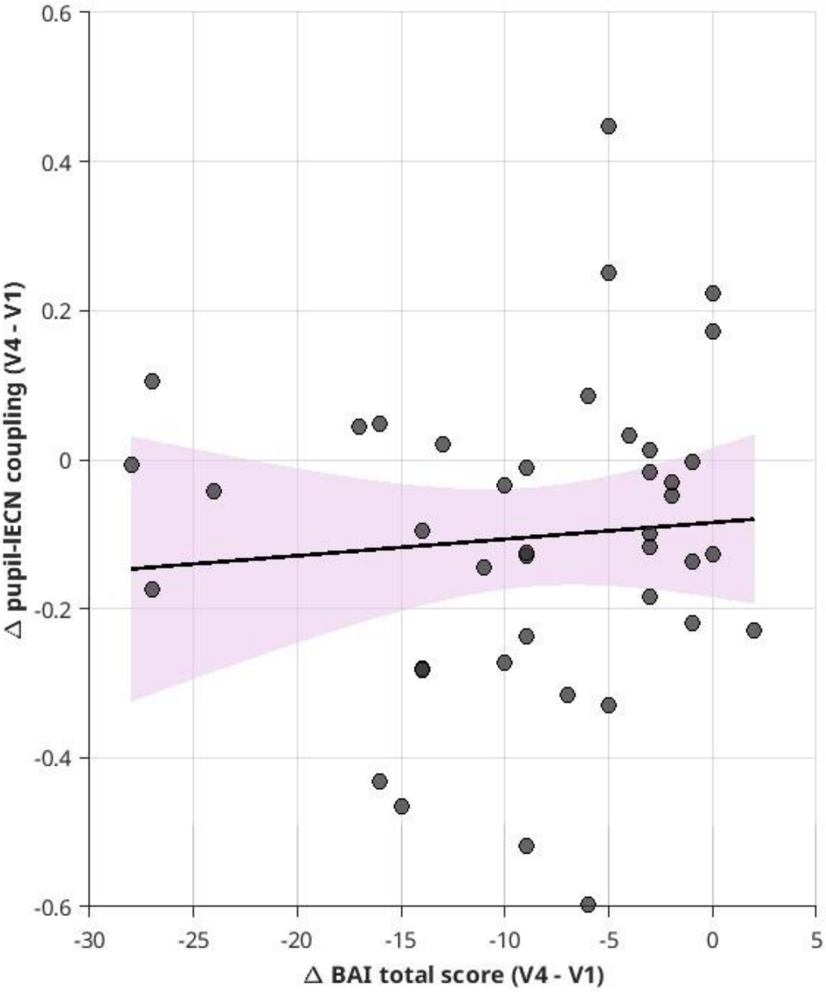


**Figure S7 Association between changes in anxiety symptoms (Beck Anxiety Inventory - BAI) and left executive control network (lECN) pupil coupling.** Scatterplot showing the association between changes in Beck Anxiety Inventory (BAI) scores and changes in pupil size-left executive control network (lECN) coupling from baseline (V1) to the post-crossover visit (V4). Each point represents one participant. The solid line represents the fitted linear regression line, and the shaded pink area indicates its 95% confidence interval. Changes in BAI scores were not significantly associated with changes in pupil–lECN coupling (Pearson's $r = 0.08$, $p = 0.60$).

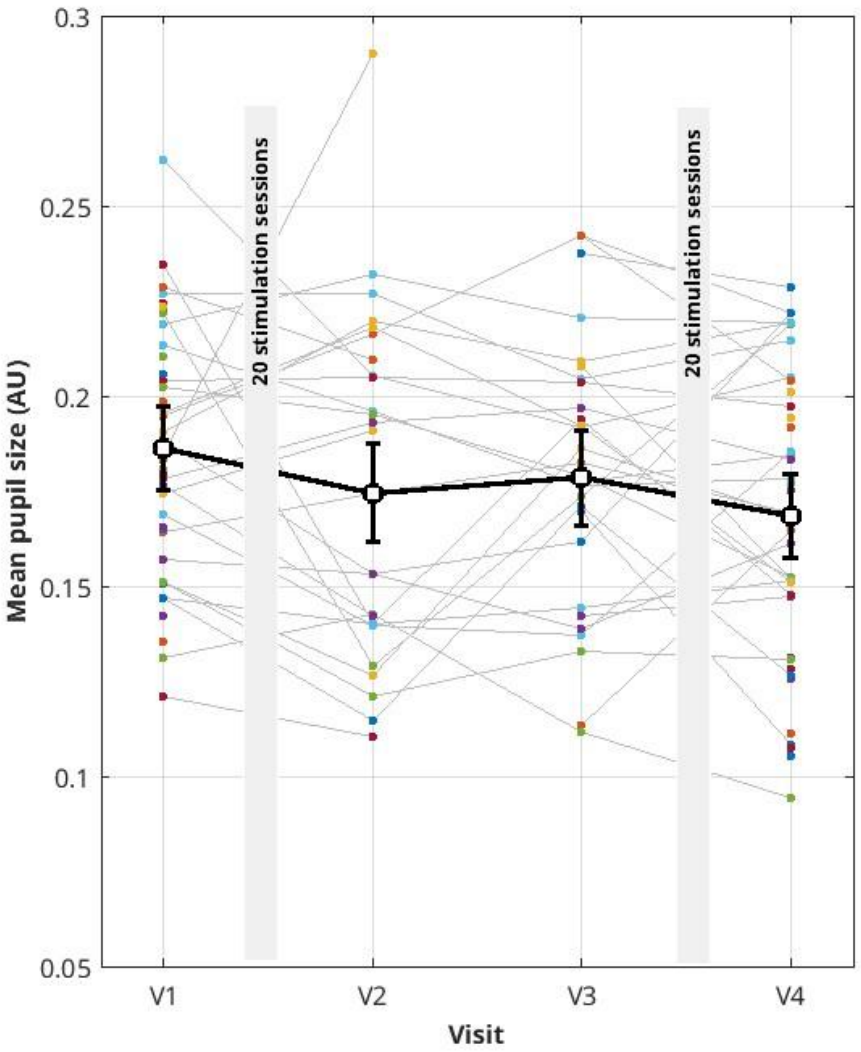


**Figure S8 Sensitivity analysis on the linear mixed-effects model of mean pupil size.** Longitudinal mean pupil size observations after exclusion of the two observations with standardized marginal residuals >3 SD, together with adjusted estimated marginal means (EMMs) and 95% confidence intervals from the refitted linear mixed-effects model. The model included visit, treatment sequence, age, and sex as fixed effects and subject as a random intercept. A nominal significant longitudinal Visit effect is outlined ($F(3,128) = 3.80$, $p = 0.01$), showing an overall decrease in mean pupil diameter across the whole treatment period. Each intervention week, comprising 20 stimulation sessions, is represented by a gray bar. Mean pupil diameter metrics are expressed in arbitrary units (AU).

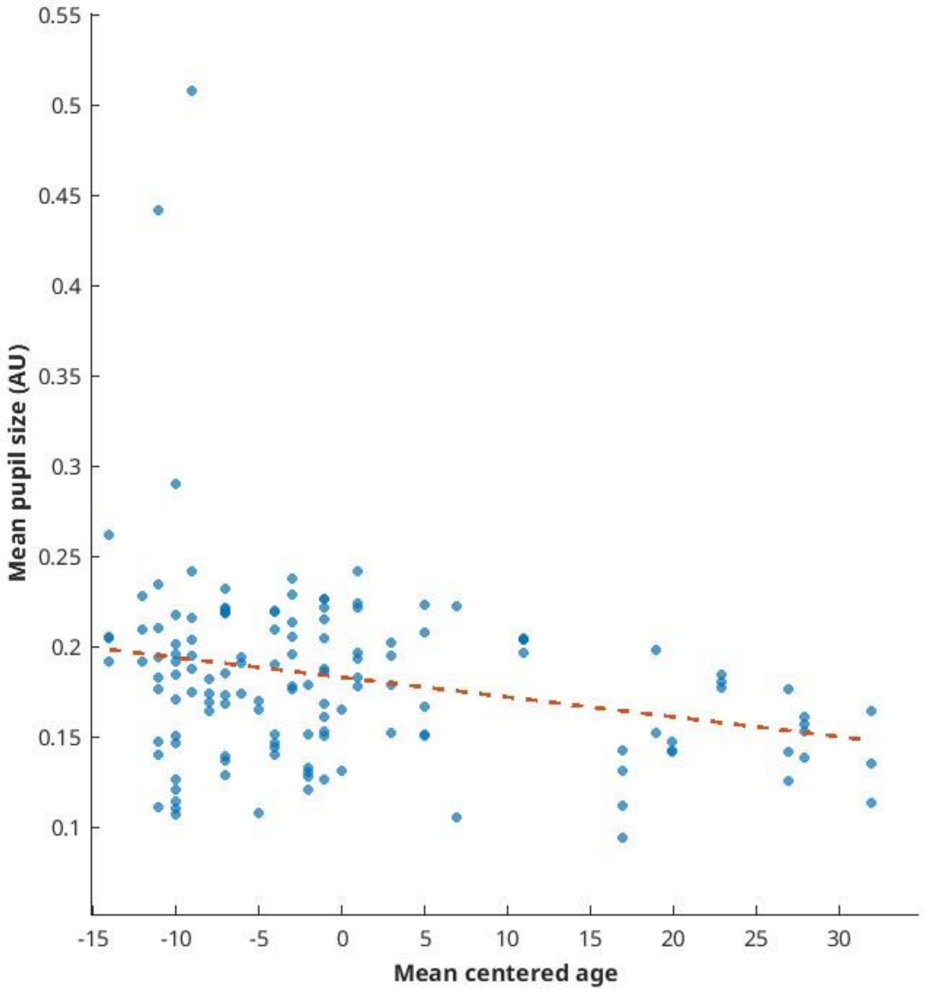


**Figure S9 Main effect of age on mean pupil size.** Scatterplot showing the association between mean centered age and mean pupil diameter across observations. The solid line represents the model-estimated age effect from the linear mixed-effects model, with predictions averaged across Visit, treatment sequence, and sex as fixed effects. Age was included as a continuous fixed-effect predictor, with subjects included as a random intercept. The corresponding omnibus test of the age effect is reported in the figure as statistically significant ($F(1,131) = 4.71$, $p = 0.03$). Mean pupil diameter metrics are reported in arbitrary units (AU).

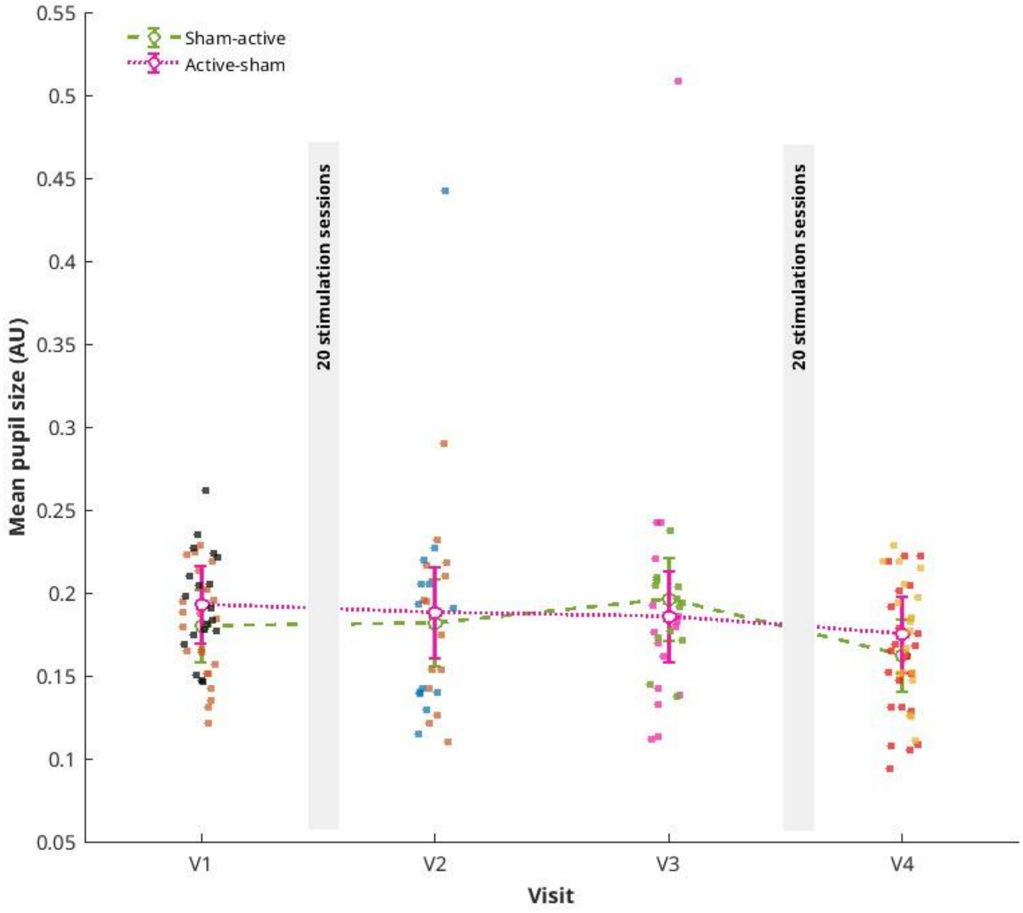


**Figure S10 Linear mixed-effects model on mean pupil size: interaction effect with treatment order.** Individual observations and model-estimated marginal means with 95% confidence intervals are shown for mean pupil diameter across study visits (V1 V2 V3 V4) in participants assigned to the sham-active and active-sham treatment sequences. Estimates are derived from a linear mixed-effects model including Visit, treatment sequence, the Visit × Sequence interaction, age, and sex as fixed effects and subject as a random intercept. The Visit × Sequence interaction was assessed using an omnibus test. Pupil size was shown to be not significantly moderated by treatment order across all visits, as no Visit x Sequence was found ($F(3,128) = 0.51$, $p = 0.67$). Each intervention week, comprising 20 stimulation sessions, is represented by a gray bar. Mean pupil diameter metrics are reported in arbitrary units (AU).

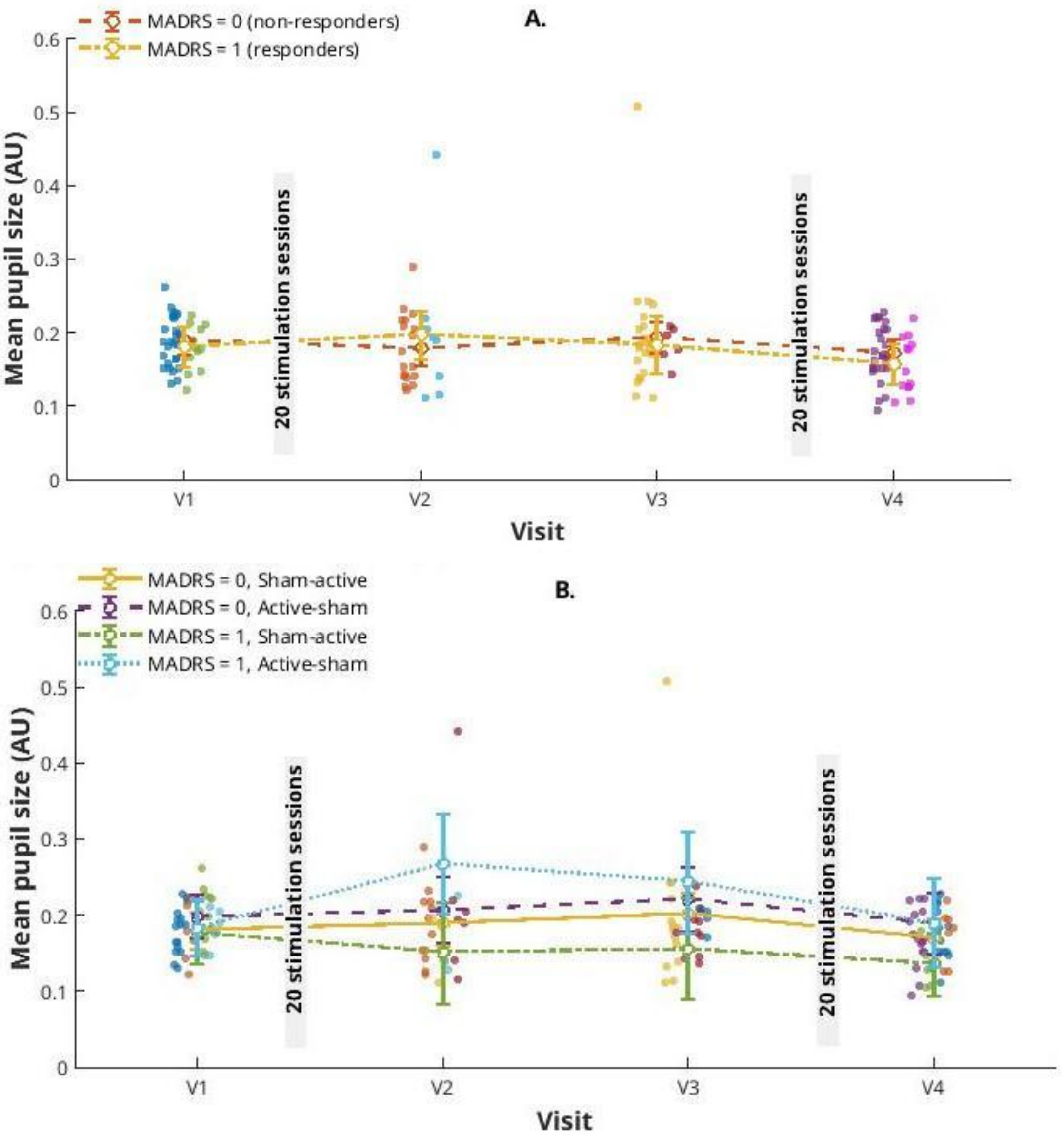


**Figure S11 Clinical moderation of MADRS treatment response on mean pupil diameter. (A)** Model A shows model-estimated marginal means and 95% confidence intervals for mean pupil size across visits (V1 V2 V3 V4) separately for participants classified as MADRS = 0 (not-responders) and MADRS = 1 (responders). **(B)** Model B further incorporates treatment sequence, showing model-estimated marginal means and 95% confidence intervals across visits for the combinations of MADRS response status and sham-active or active-sham treatment sequence. Individual observations are overlaid in both panels. Model A included Visit, MADRS status, their interaction. Model B additionally included treatment sequence and the corresponding Visit × Sequence × MADRS interaction. In both models, age and sex are treated as fixed effects and subject as random intercept. The respective interaction terms were assessed using omnibus tests. Pupil size was not significantly associated with MADRS responder status (Panel A; $F(3, 127) = 0.84$, $p = 0.47$). Similarly, the three-way interaction involving visit, MADRS responder status, and treatment order was not statistically significant (Panel B; $F(3, 120) = 1.95$, $p = 0.12$). Each intervention week, comprising 20 stimulation sessions, is represented by a gray bar. Mean pupil diameter is reported in arbitrary units (AU).

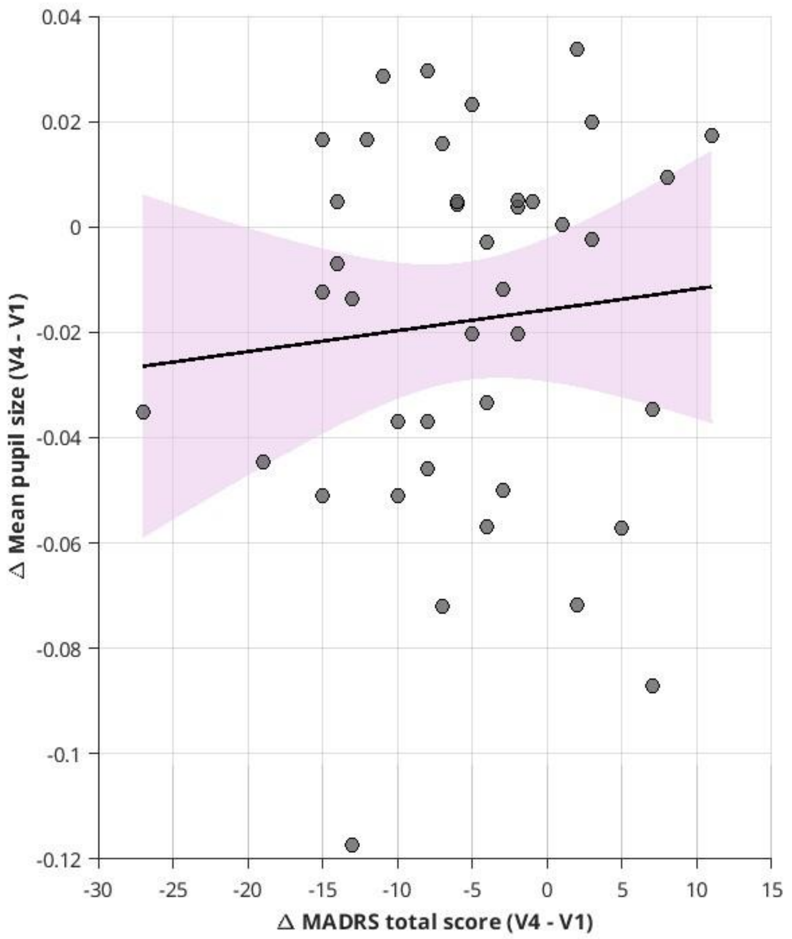


**Figure S12 Association between mean pupil size and depressive symptoms (Montgomery-Åsberg Depression Rating Scale-MADRS) from baseline (V1) to post-treatment visit (V4).** Scatterplot showing the association between V1–V4 changes in mean pupil diameter and Montgomery-Åsberg Depression Rating Scale-MADRS (MADRS) total scores, with each point representing one participant. The solid line represents the fitted linear regression line, and the shaded pink area indicates its 95% confidence interval. No significant correlation between total MADRS scores changes and mean pupil size changes at V1-V4 was found ($r = 0.09$, $p = 0.57$). Mean pupil diameter metrics are reported in arbitrary units (AU).

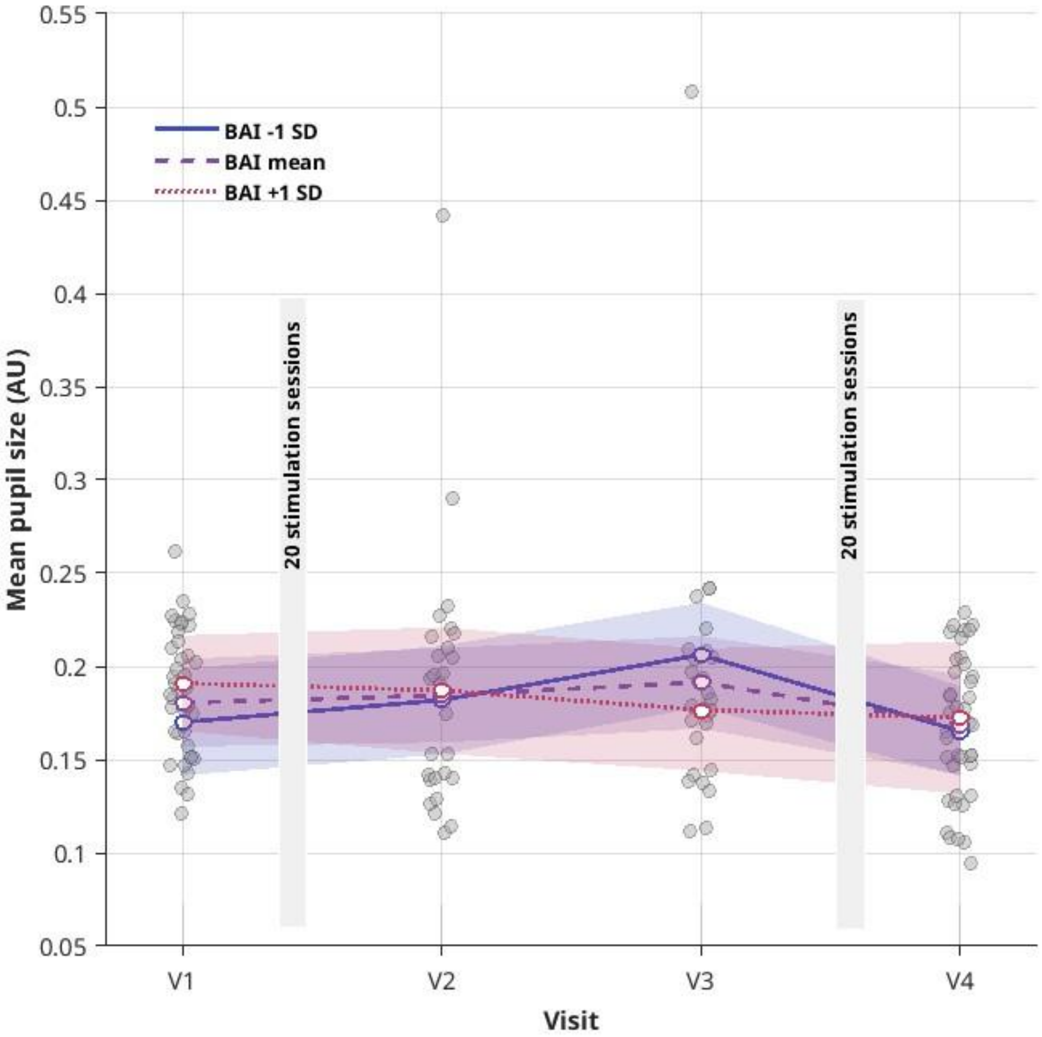


**Figure S13 Longitudinal mean pupil size changes as a function of Beck Anxiety Inventory (BAI) symptom level.** Observed mean pupil diameter values are shown as individual points at each visit, with jitter applied horizontally for visualization. Colored lines and points represent estimated marginal means (EMMs) from the longitudinal linear mixed-effects model at the mean Beck Anxiety Inventory (BAI) score and at ±1 SD from the mean BAI score; shaded areas indicate the corresponding 95% confidence intervals. The EMMs represent model-based predictions for representative levels of the continuous BAI moderator. The model included Visit × BAI interaction, treatment sequence, age, and sex as fixed effects, with a subject-level random intercept. No significant longitudinal effect of BAI total scores was found moderating pupil size across all visits ($F(3,127) = 1.99$, $p = 0.11$). Each intervention week, comprising 20 stimulation sessions, is represented by a gray bar. Mean pupil diameter metrics are reported in arbitrary units (AU).
